# Proteogenomics captures a midlife biological state of Alzheimer’s disease susceptibility

**DOI:** 10.64898/2026.10.22.26366029

**Authors:** Andrew S. Perry, Hailey A. Adegboye, Xavier Bledsoe, Derek Archer, Anshul Tiwari, Phillip Lin, Eric Farber-Eger, B. Lakshitha A. Perera, Shilin Zhao, Quanhu Sheng, Siyuan Ma, Elizabeth Hutchins, Panpan Zhang, Madeline Wood Alexander, Rebecca Sale, Yike Chen, Laura Colangelo, Amber Bahorik, Kahraman Tanriverdi, Antonina Risitano, Jane E. Freedman, Matthew Nayor, Saumya Das, Dandan Liu, Kimberly R. Pechman, Niranjana Shashikumar, Corey Bolton, L. Taylor Davis, Bennett A. Landman, Lifang Hou, Carlos Cruchaga, Rowan Saloner, Kristine Yaffe, Quinn S. Wells, Timothy J. Hohman, Angela L. Jefferson, Kendall van Keuren-Jensen, Ravi V. Shah, Eric R. Gamazon

## Abstract

Alzheimer’s disease (AD) develops over decades, but biological processes underlying susceptibility during midlife remain poorly understood. Using multimodal neuroimaging and plasma proteomics in UK Biobank participants, we identified circulating proteomic signatures of brain structure and organization. This ‘neurostructural proteome’ predicted future cognitive decline, incident dementia, and later-life AD pathology across independent cohorts spanning two decades of follow-up. Genetic and single-nucleus transcriptomic analyses demonstrated convergence of the neurostructural proteome and AD genetic architecture, highlighting enrichment in astrocyte, microglia, and endothelial pathways beyond the APOE region. Collectively, these findings identify a circulating neurostructural proteome detectable in midlife that captures molecular processes associated with future AD susceptibility and provide a framework for uncovering an early biological landscape of disease susceptibility that precedes conventional biomarkers and clinical onset.

## INTRODUCTION

Alzheimer’s disease (AD) will impact 153 million individuals by 2050^1^. Modern approaches to AD detection using fluid biomarkers target underlying neuropathology (e.g., phosphorylated tau-217 [pTau-217]^2–4^, amyloid-beta [Aβ]^5^)^6^. Nevertheless, plasma pTau-217 is rarely elevated in individuals under age 60 years without established cognitive impairment or genetic AD (<4% in some studies^7^). Recent data suggest that easily accessible circulating neuropathological biomarkers may have limited positive predictive value for the earliest detection of AD, with a potential benefit of combination biomarker approaches to stratify early susceptibility^8^. Midlife (≈40-55 years) is a critical period for the emergence of biological factors relevant to brain aging^9^, during which metabolic-inflammatory processes (e.g., obesity, insulin resistance, inflammation) may potentiate later cognitive decline and AD onset^10–12^. Given the value of addressing risk at its pre-symptomatic stage to prevent disease in other contexts^13,14^, examining the midlife phase as a critical, upstream phase of AD susceptibility through neurostructural signatures and brain-specific cellular programs may illuminate upstream pathobiological processes central to potential intervention and risk assessment before substantial neurodegeneration occurs.

Here, we hypothesized that variation in brain architecture could serve as a biological anchor for identifying molecular correlates of neurostructural variation linked to AD susceptibility that precede established neuropathology. By integrating brain magnetic resonance imaging (MRI) and circulating proteomics in >8,000 UK Biobank participants, we decomposed 256 brain MRI-defined imaging traits into three composite neurostructural phenotypes reflecting grey matter, white matter, and cerebrovascular-neurodegenerative integrity. This circulating “neurostructural proteome” differed significantly in AD among >9,000 individuals in the Global Neurodegeneration Proteomic Consortium, with genetic localization to brain regions and white matter tracts implicated in AD in >33,000 individuals. Multi-protein scores reflecting neurostructural phenotypes were additive to AD genetic liability in prognostication and improved discrimination of AD susceptibility in an independent UK Biobank sample. We quantified the multi-protein scores from the fifth decade of life (40s) onward in two cohorts—the Coronary Artery Risk Development in Young Adults (CARDIA) and Vanderbilt Memory and Aging Project (VMAP)—and found that lower multi-protein scores were associated with accelerated cognitive decline and greater grey matter atrophy. Higher multi-protein scores in the 5^th^ decade of life (40s) associated with a more beneficial plasma biomarker profile of AD while favorable longitudinal changes in multi-protein scores over two decades were associated with better cognitive performance. Finally, using genetic models of single-nucleus RNA expression in brain, we identified cell type-resolved differential gene expression in AD and cell type enrichment of the neurostructural proteome within AD (111,326 cases and 677,663 controls)— but not Parkinson’s disease (67,834 cases and 2,458,063 controls)—genetic risk loci.

Collectively, these findings identify a circulating neurostructural proteome in midlife that reflects cell type-dependent AD pathobiology in the brain and captures molecular processes that precede AD biomarker positivity, motivating a shift toward studying and intervening on *midlife biology*, where pathogenic trajectories are observable and preventive strategies may be most effective.

**Figure 1.**
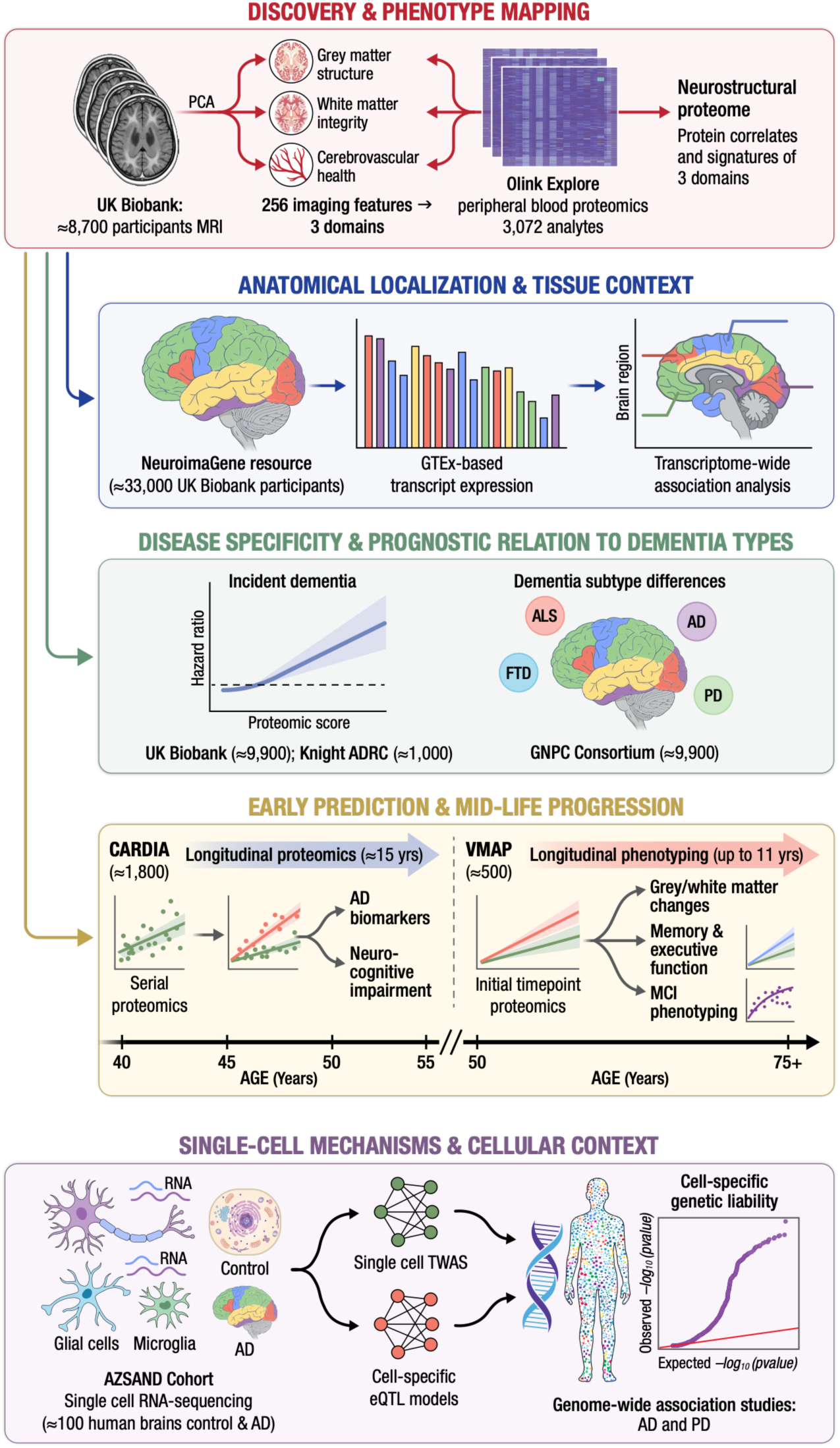
Study overview | Graphical abstract.

## RESULTS

### Circulating proteomics of composite brain MRI-derived neurostructural phenotypes reflect metabolic mechanisms in AD

To characterize the relationships between circulating, plasma-based proteins and neurostructural phenotypes, we studied 8,711 participants with plasma proteomics data (Olink Explore 3072) who underwent brain MRI in UK Biobank (age 54±8 years, 54% female; **Supplemental Table 1**). We decomposed 256 brain imaging-derived phenotypes (IDPs) into three principal components, together (PC1, PC2, PC3), accounting for ≈30% of the variance in the brain MRI phenome (**Figures 2A-B, Supplemental Table 2**). The first 3 PCs were retained because they captured interpretable, principal axes of variation while substantially reducing dimensionality (**Supplemental Figure 1**). Grey matter structural measures, including FreeSurfer grey matter volumes and cortical thickness measures, loaded into PC1, labeled “grey matter structure”; lower PC1 values indicate loss of grey matter tissue consistent with neurodegeneration. PC2 was loaded on white matter microstructural measures, including diffusion tensor imaging (DTI) fractional anisotropy (DTI-FA) and DTI intracellular volume fraction (DTI-ICVF), and was labeled “white matter integrity”; lower PC2 values indicate loss of white matter microstructural integrity and reduced neurite density. PC3 combined grey matter volume, white matter microstructural integrity, and white matter macrostructural injury measures, including functional Magnetic Resonance Imaging of the Brain’s (FMRIB) Integrated Registration and Segmentation Tool (FIRST) subcortical grey matter volumes, DTI isotropic volume fraction of isotropic diffusion (DTI-ISOVF), DTI orientation dispersion index (DTI-ODI), and white matter hyperintensity (WMH) volume, and was labeled “cerebrovascular-neurodegenerative integrity” (referred to hereafter as “cerebrovascular health” for brevity). PC3 was positively correlated with subcortical grey matter volumes and DTI-ODI and negatively correlated with DTI-ISOVF and WMH volume, such that lower PC3 values reflect tissue profiles consistent with cerebrovascular pathology.

**Figure 2.**
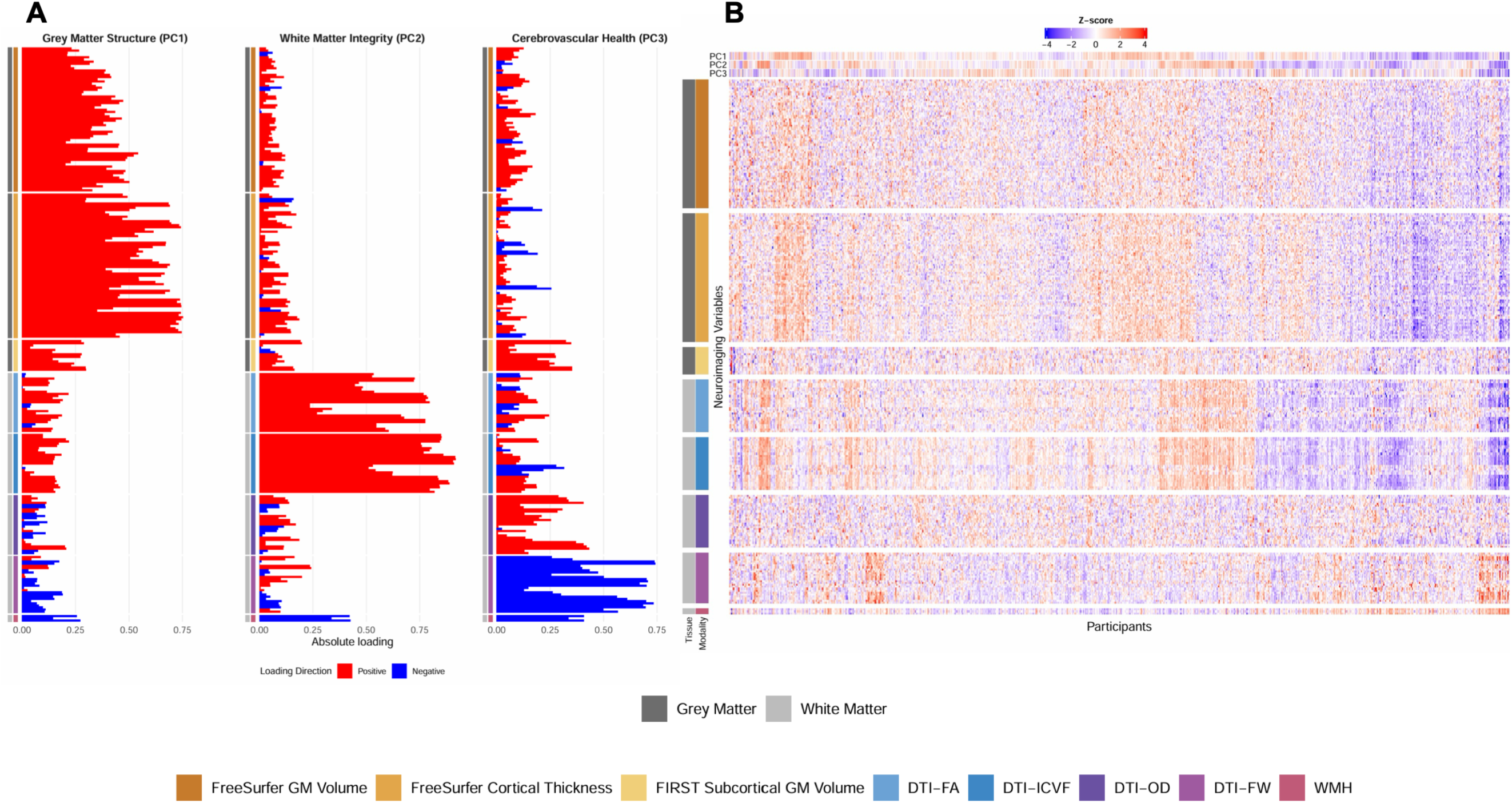
Principal components of neuroimaging data. **(A)** Principal component analysis identified 3 PCs (from 256 brain MRI imaging-derived phenotypes) that capture ≈30% of the variance in neuroimaging data. Visualized are the principal component loadings for each of the 3 PCs. **(B)** Heatmap of the brain imaging data with annotations for the PCs. The main body of the heatmap shows standardized values, and the annotation bars at the top represent the participants related PC score, derived from the loadings shown in panel A. DTI = diffusion tensor imaging, FA = fractional anisotropy, ICVF = intracellular volume fraction, ODI = orientation dispersion index, FW = free water, WMH = white matter hyperintensity, FIRST = Functional Magnetic Resonance Imaging of the Brain’s Integrated Registration and Segmentation Tool, GM = grey matter.

In age- and sex-adjusted models, we identified a broad set of proteins associated with each IDP-derived PC, with the most associations observed for PC3 (**Figure 3A**, **Supplemental Table 3**). Across the three neurostructural PCs, 1,172 unique proteins (of 2,920 tested) were significantly associated with at least one PC at FDR<5% (full list in **Supplemental Table 3**), constituting the “neurostructural proteome;” 172 proteins were associated with two or more PCs. Effect sizes were robust to further adjustment for race/ethnicity, BMI, education, smoking, Townsend Deprivation Index, and *APOE*-ε4 status (Pearson R=0.92 across all models; **Supplemental Table 4**). Proteins associated with all 3 PCs included GFAP (an established plasma biomarker for dementia^15^ involved in astrogliosis) along with markers of neuroinflammation (IL1RL2^16^, LGMN^17^, REN^18^), neuronal injury (HGF^19^, KLK6^20^, SMPD1^21^), Aβ processing (TIMP1^22^, MDK^23^, KLK6^20^, PTPRR^24^), and cholinergic signaling (BCHE^25^), among others. Proteins identified in UK Biobank were differentially abundant in the Global Neurodegeneration Proteomic Consortium (GNPC; n=9,972, age 70±12 years, 56% female) across a range of neurodegenerative conditions, including AD, amyotrophic lateral sclerosis (ALS), frontotemporal dementia (FTD), and Parkinson’s disease (PD) with the <u>largest</u> <u>differences observed between AD and cognitively normal</u> (**Figure 3B**, **Figure 4, Supplemental Table 5**). Pathway analysis across Gene Ontology Biological Process (GO BP), Kyoto Encyclopedia of Genes and Genomes (KEGG), Reactome, and WikiPathway databases, using targets with age- and sex-adjusted FDR<5% in UK Biobank, identified extracellular matrix organization and disassembly, cytokine-cytokine interactions, lipid transport, and neuronal injury from hyperglycemia (**Supplemental Table 6**). While many targets have been previously implicated in the biology of dementia, novel findings included CEACAM16^26^ (related to cochlear function, of interest due to the association between hearing loss and dementia) and BPIFB2^27^ (involved in innate immunity and lipid transfer).

**Figure 3.**
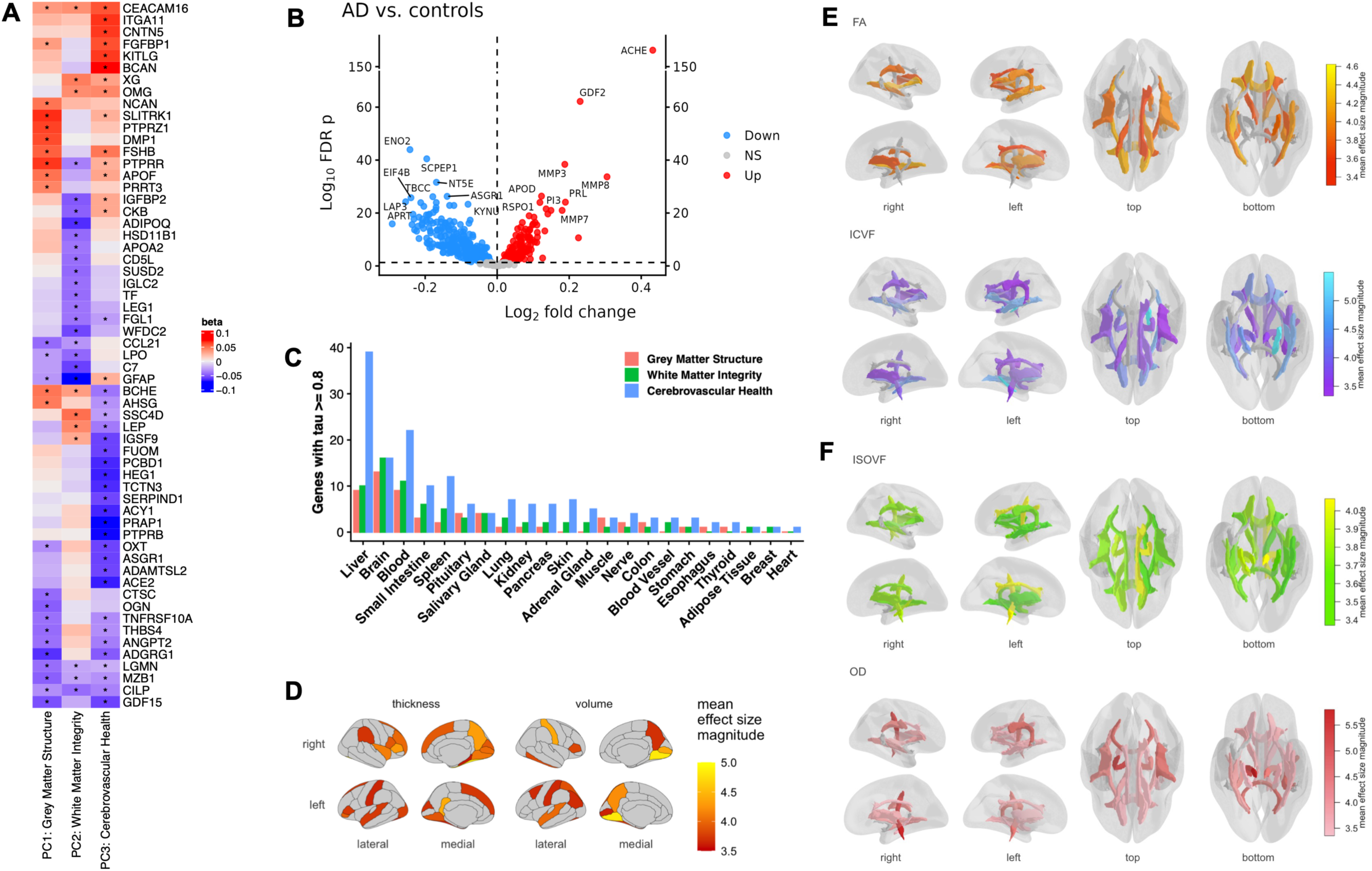
Proteomic profiling of neurostructural phenotypes. **(A)** Heatmap of beta coefficients from UKB linear models of plasma proteomic measures and neurostructural PCs. For visualization, we selected the top 20 proteins for each PC, ranked by absolute value of beta coefficient. Asterisk indicates FDR<0.05. **(B)** Volcano plots illustrating differential abundance of proteins related to neurostructural PCs at an FDR<5% across AD and cognitively normal in GNPC. **(C)** Tissue of origin using GTEx-derived tau index for proteins related to neurostructural PCs at an FDR<5%. Tau ≥0.8 is considered to have a high-degree of tissue specificity. **(D)** Grey matter correlates of PC1 genes. We tested the genes encoding the proteins associated with PC1 for associations with grey matter regions. For each region we aggregate the GReX of all Bonferroni significant genes across both thickness and volume, coloring regions according to the mean effect size magnitude. Regions are depicted according to the Desikan cortical atlas. **(E)** White Matter correlates of PC2 genes. We tested the genes encoding the proteins associated with PC2 for associations with white matter tractography IDPs. For each IDP we aggregate the GReX of all Bonferroni significant genes (Bonferroni-corrected P<0.05) across both FA and ICVF, coloring regions according to the mean effect size magnitude. Tract IDPs are depicted according to the JHU tractography atlas. **(F)** White Matter correlates of PC3 genes. We tested the genes encoding the proteins associated with PC3 for associations with white matter tractography IDPs. For each IDP we aggregate the GReX of all Bonferroni significant genes across both ISOVF and OD, coloring regions according to the mean effect size magnitude. Tract IDPs are depicted according to the JHU tractography atlas. BMI = body mass index; TDI = Townsend Deprivation Index; AD = Alzheimer’s disease; ALS = amyotrophic lateral sclerosis; FTD = frontotemporal dementia; PD = Parkinson’s disease.

**Figure 4.**
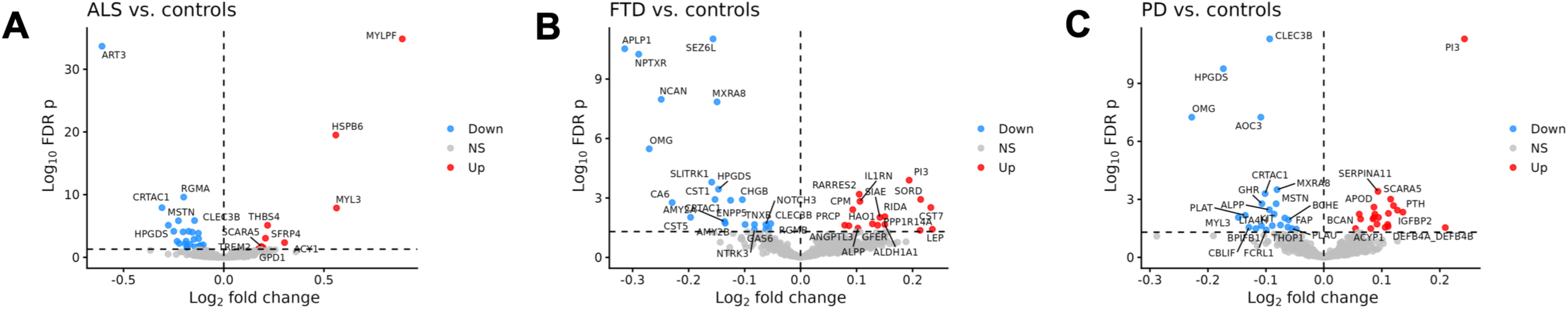
Proteomic markers of neurostructural phenotypes across neurodegenerative diseases. **(A, B, C)** Volcano plots illustrating differential abundance of proteins related to neurostructural PCs at an FDR<5% across neurodegenerative phenotypes in GNPC. ALS = amyotrophic lateral sclerosis; FTD = frontotemporal dementia; PD = Parkinson’s Disease.

### Tissue and brain anatomical localization of the neurostructural proteome

A significant fraction of the neurostructural proteome was enriched in liver, brain, and blood, as determined by the tau (τ) tissue-specificity index, which assigns each gene an enrichment score reflecting how concentrated its expression is in a given tissue relative to others (see **Methods**; **Figure 3C**), suggesting biological processes that span peripheral metabolism and central nervous system function implicated in AD^28,29^. Integration with the NeuroimaGene resource^28^ further localized these molecular signals to neuroanatomical regions implicated by the neurostructural proteome, providing convergent evidence that systemic molecular variation maps onto specific structural substrates of the human brain. The NeuroimaGene resource—derived from 33,000 UK Biobank participants and GTEx-based genetic models of brain region-specific transcript expression—allowed us to examine whether gene transcripts for the neurostructural proteome were associated with specific anatomical features via genetically regulated gene expression (GReX). To identify the neuroanatomical substrates of the neurostructural proteome, we tested tissue-specific GReX models for proteins associated with each PC using anatomically matched imaging-derived phenotypes. Specifically, proteins linked to grey matter structure (PC1) were evaluated against FreeSurfer-derived cortical thickness and grey matter volume measures, proteins linked to white matter integrity (PC2) against DTI-FA and DTI-ICVF metrics, and proteins linked to cerebrovascular health (PC3) against DTI-OD and DTI-ISOVF measures.

The genetically regulated expression of genes encoding 21 (11.0%) proteins associated with PC1 was associated with grey matter volume and cortical thickness measures (Bonferroni-corrected P<0.05, **Figure 3D**, **Supplemental Table 7**). The top association for PC1 by effect size implicated GReX of *NADK* (implicated in AD pathogenesis^30^) with the volume of the right postcentral gyrus. GReX of 12 (4.9%) proteins associated with PC2 was associated with DTI-FA and DTI-ICVF measures (Bonferroni-corrected P<0.05, **Figure 3E**, **Supplemental Table 8**), the top associations for PC2 all involved GReX of *NEFL*, a sensitive biomarker of axonal damage and AD^8^, and eight DTI-ICVF IDPs capturing white matter microstructure and axonal integrity, providing biological concordance between molecular and neuroimaging markers of axonal injury. GReX of 116 (12.6%) proteins associated with PC3 demonstrated associations with DTI-ISOVF and DTI-OD measures (Bonferroni-corrected P<0.05, **Figure 3F**, **Supplemental Table 9**). For PC3, GReX of *KHK* was strongly associated with DTI-OD in the superior longitudinal fasciculus. Ketohexokinase (KHK, fructose metabolic pathway in microglia) has demonstrated metabolic effects on progressive cognitive dysfunction in diabetic model systems^31^ and has been proposed as a potential target in AD^32^.

Overall, the NeuroimaGene results provide genetically determined transcriptional-level support that the neurostructural proteome reflects both grey matter regions previously shown to have decreased perfusion in individuals at-risk for AD^29^ and white matter tracts critical for cognition^30^ (e.g., executive function, language), pointing to specific structural and metabolic mechanisms of AD risk (e.g., *NEFL*^8^, *KHK*, *NADK*; **Supplemental Table 10**).

### Multi-protein scores of neurostructural phenotypes identify risk of long-term AD independent of genetic liability and standard clinical risk factors

Using independent derivation and validation samples from UKB, we constructed and validated least absolute shrinkage and selection operator (LASSO) models of neurostructural PCs using only proteins that were significant in age- and sex-adjusted models for the relevant PC at 5% FDR (**Figure 5A**, **Supplemental Table 11**). For example, the LASSO model of PC1 was constructed from the 191 proteins related to PC1 in age- and sex-adjusted linear models at FDR<5% (**Supplemental Table 3**). Resulting multi-protein scores for each composite neurostructural PC (number of proteins selected for PC1=84, PC2=102, PC3=137) were correlated with their parent PC phenotypes (across derivation and validation samples, Pearson r 0.37-0.56, p<0.0001; **Supplemental Figure 2**). Multi-protein scores were negatively associated with age (Pearson r −0.67 to −0.57, p<0.0001) and differed by sex, but not by *APOE*-ε4 status (**Figure 5B**, **Figures 6A-C**). Multi-protein scores restricted to the top 30 coefficients (ranked by absolute value of LASSO coefficient) correlated strongly with the original full-length scores (Pearson r 0.94-0.96, p<0.0001, **Supplemental Figure 3A**), suggesting effect sizes are largely preserved when reduced to a dimension consistent with current commercially available proteomics platforms.

**Figure 5.**
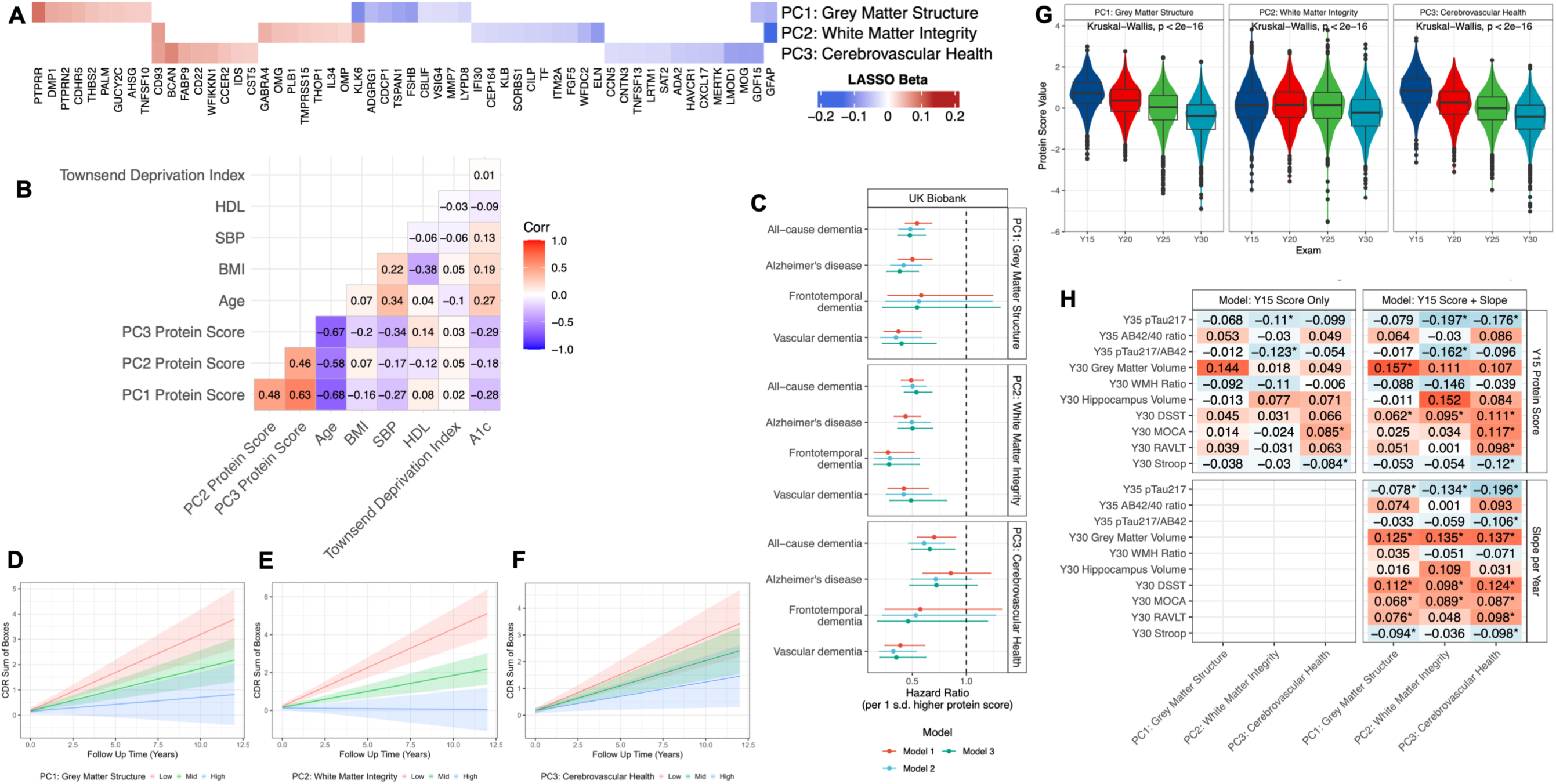
Multi-protein scores of neurostructural phenotypes are associated with dementia. **(A)** Heatmap of beta coefficients from LASSO models of neurostructural PCs. For visualization, the top 20 proteins ranked by absolute value of beta coefficient are shown. **(B)** Pearson correlation heatmap between multi-protein scores of neurostructural phenotypes with key clinical risk factors. **(C)** Forest plot representing Cox model results for each multi-protein score and its association with all-cause dementia and sub-types of dementia in UK Biobank. Error bars represent 95% confidence intervals of the hazard ratio. Colors represent various sets of covariate adjustments: Model 1: age, sex; Model 2: Model 1 + race, body mass index, smoking status, education level, APOE-ε4 status; Model 3: Model 2 + systolic blood pressure, high-density lipoprotein, hemoglobin A1c, Townsend Deprivation Index. **(D, E, F)** Longitudinal clinical trajectories, assessed via the Clinical Dementia Rating Sum of Boxes Score across tertiles of protein scores in VMAP. **(G)** Distribution of multi-protein risk scores across Exam Years 15-30 in CARDIA demonstrate an overall decline in score. **(H)** Heatmap of beta coefficients from linear models of cognitive outcomes in CARDIA at Year 30 and neurodegenerative biomarkers at Year 35 as a function of Year 15 multi-protein risk scores and estimated slope from Year 15 to Year 30. Models adjusted for age, sex, race, Year 30 education, APOE-ε4 status. Asterisk indicates FDR<0.05. AD = Alzheimer’s disease; ALS = amyotrophic lateral sclerosis; FTD = frontotemporal dementia; PD = Parkinson’s disease; CU = cognitively unimpaired; MCI = mild cognitive impairment; CDR = Clinical Dementia Rating; WMH Ratio = white matter hyperintensity ratio.

**Figure 6.**
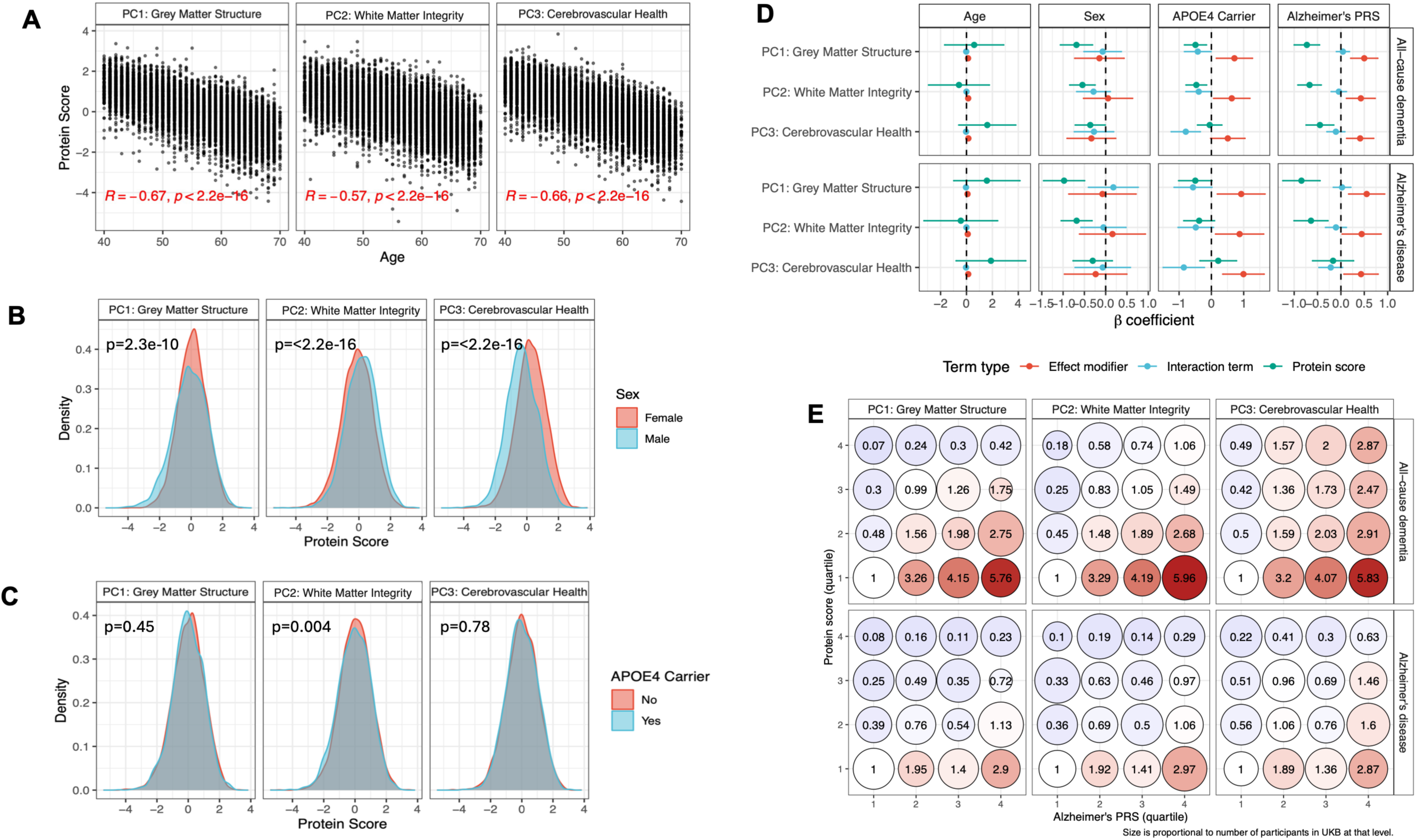
Synergistic effects of multi-protein scores with demographic and genetic factors. **(A)** Scatterplots of multi-protein scores relations with age. **(B, C)** Density histograms comparing multi-protein score distribution across sex and *APOE*-ε4 status. Reported p-values are from Wilcoxon tests. **(D)** Forest plot of Cox models of all-cause dementia and Alzheimer’s disease including interaction terms between the multi-protein scores and age, sex, *APOE*-ε4 status, and polygenic risk score of Alzheimer’s disease**. (E)** Balloon plots showing hazard ratios for outcomes of all-cause dementia (top row) and Alzheimer’s disease (bottom row). Cell color is proportional to the hazard ratio. HR are set relative to the bottom quartile of both multi-protein score and PRS.

In 9,973 UKB participants without dementia at study entry, we observed 114 cases of all-cause dementia, 60 cases of AD, 25 cases of vascular dementia, and 9 cases of FTD over a median 13.2 years (25^th^-75^th^ percentile: 11.7-14.1 years). Dementia subtypes were defined by ICD codes (see **Methods**), which likely reflects clinical presentations rather than biomarker defined cases. In Cox regressions, higher, circulating, multi-protein scores for all three neurostructural PCs were associated with substantially lower risk of all-cause dementia (age/sex-adjusted hazard ratio [HR] and 95% CI: PC1 Grey Matter Structure 0.54 [0.44-0.68], PC2 White Matter Integrity 0.49 [0.40-0.60], PC3 Cerebrovascular Health 0.70 [0.55-0.90]; all p≤0.005; **Figure 5C, Supplemental Table 12**). Similar effect sizes and directionality were observed for AD, vascular dementia, and FTD (with power limited due to few events). Of note, multi-protein score associations were robust to further adjustment for race, body mass index (BMI), education, smoking status, Townsend deprivation index (TDI), *APOE*-ε4 status, systolic blood pressure, high-density lipoprotein, and hemoglobin A1c (**Supplemental Table 12**). The multi-protein score for grey matter structure (PC1), but not other neurostructural PCs, significantly improved risk discrimination for all-cause dementia beyond established risk factors (C-index 0.835 to 0.850, p=0.04; **Supplemental Table 13**), indicating that the multi-protein scores contain information not fully captured by established risk factors. We did not observe significant interactions between multi-protein scores and age, sex, *APOE*-ε4 status, or AD polygenic risk score (PRS) on dementia risk (**Figure 6D, Supplemental Table 14**). We did observe independent, additive effects of each multi-protein score and AD PRS in Cox regression for all-cause dementia and AD, with the additive effects in the highest adverse quartile of genetic and proteomic risk (**Figure 6E**; combined genetic and proteomic HR for AD ≈2.9 across all 3 multi-protein scores) and protective effects of proteomics on genetic liability (**Figure 6E, Supplemental Table 15**). Association effect sizes were preserved in multi-protein scores composed of the top 30 proteins ranked by absolute coefficient value (**Supplemental Figure 3B**).

### Multi-protein scores of neurostructural phenotypes differentiate AD diagnosis and mild cognitive impairment across four independent cohorts

To assess the broader diagnostic and prognostic potential of these multi-protein scores, we examined four independent cohorts across the life-course: (1) Coronary Artery Risk Development in Young Adults (CARDIA, serial proteomics from ages ≈40 to ≈55, 58% female, with cognitive health and MRI at age 55); (2) Vanderbilt Memory and Aging Project (VMAP; age ≈65, 53% female, with a median 4 years of serial cognitive assessments); (3) GNPC (cross-sectional assessment of dementia type and cognitively normal participants without dementia, age ≈70, 58% female); (4) Knight Alzheimer’s Disease Research Center (ADRC; age ≈75, 59% female, proteomics with incident dementia). Knight ADRC and GNPC proteomics data were generated using aptamer-based platforms (Somalogic), with excellent re-calibration of LASSO models using proteins common across Olink and Somalogic platforms (see **Methods**; Pearson r 0.85-0.97, p<0.0001, **Supplemental Figure 3C, Supplemental Table 16**, association with events in **Supplemental Figure 3D**, **Supplemental Table 12**).

In 2,835 GNPC participants (age ≈69, 58% female) with complete data to generate the 3 multi-protein scores (639 AD, 79 ALS, 66 FTD, 200 PD, and 1,851 cognitively normal), we observed lower PC1 (grey matter structure) and PC2 (white matter integrity) scores in participants with AD, after adjustment for age, sex, and site (**Figure 7A-C, Supplemental Table 17**). Differences across ALS, FTD, and PD were inconsistent, potentially reflecting the limited number of cases in GNPC. Similarly, in the VMAP Legacy and Expansion cohorts (age 66±9, 53% female), multi-protein scores differentiated early stages of cognitive impairment, prior to onset of clinical dementia. In both age, sex, cohort-adjusted and fully adjusted models (race/ethnicity, *APOE*-ε4 status, smoking status, BMI, area deprivation index), VMAP participants with mild cognitive impairment (MCI, n=62) exhibited lower white matter integrity (PC2) and cerebrovascular health (PC3) multi-protein scores relative to cognitively unimpaired participants (n=484) with moderate standardized differences between MCI and cognitively unimpaired individuals (0.23-0.40; **Figure 7D-F**, **Supplemental Table 18**).

**Figure 7.**
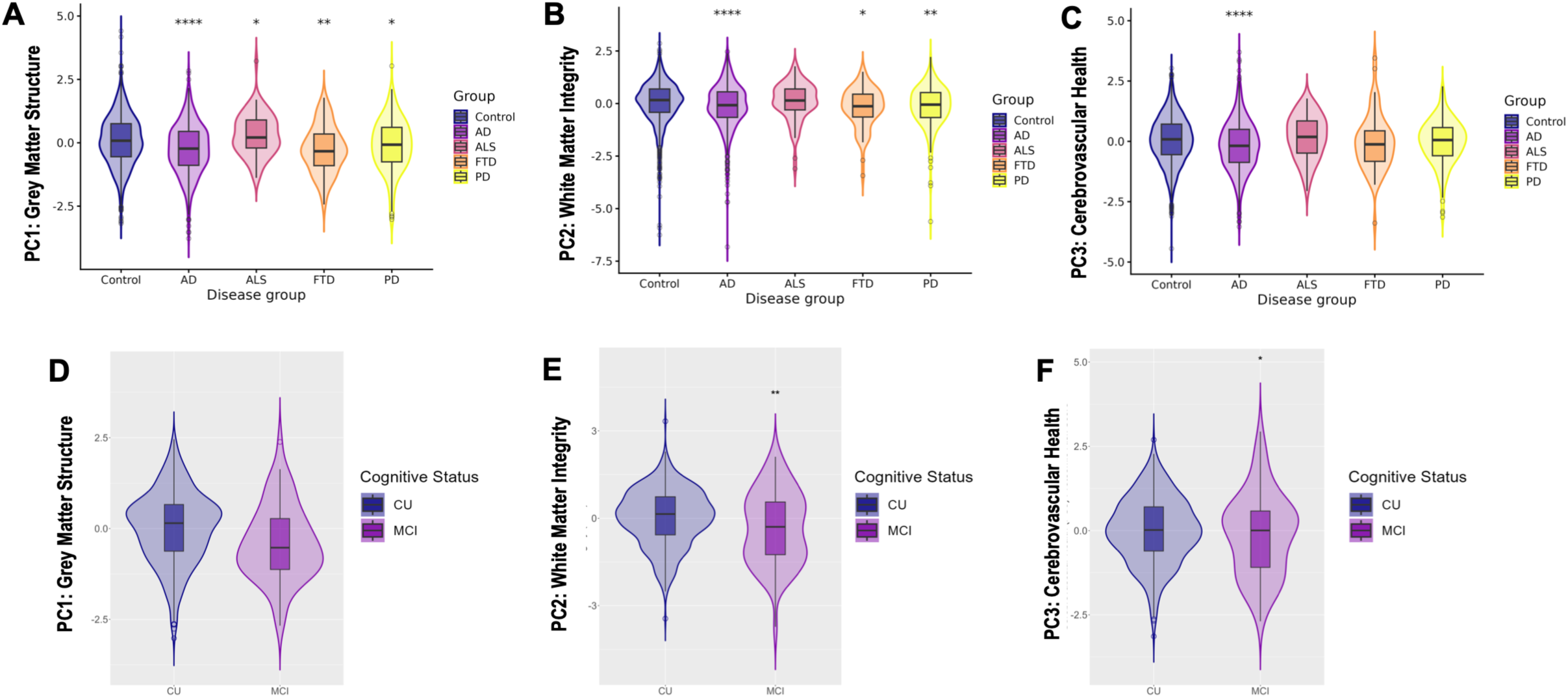
Differences in multi-protein scores across neurodegenerative phenotypes. **(A, B, C)** Comparison of multi-protein scores across neurodegenerative phenotypes in GNPC (*p<.05, **p<.01, ***p<.001, ****p<.0001). **(D, E, F)** Comparison of multi-protein scores between mild cognitive impairment and cognitively normal in VMAP (*p<0.05, **p<0.01). AD = Alzheimer’s disease; ALS = amyotrophic lateral sclerosis; FTD = frontotemporal dementia; PD = Parkinson’s disease; CU = cognitively unimpaired; MCI = mild cognitive impairment; CDR = Clinical Dementia Rating.

### Multi-protein scores predict longitudinal changes in cognitive function, brain structure, and clinical dementia scores in an AD risk population

To understand the implications of the multi-protein scores on longitudinal measures of cognition, we examined their relationship to cognitive trajectories in VMAP participants with up to 11 years of follow-up. Participants were free of clinical dementia at study entry and were examined every 1.5-2 years with clinical interview, neuropsychological assessment, and brain MRI (see **Methods**). In linear mixed effects models adjusted for age, sex, and cohort (Legacy or Expansion), lower baseline values of all multi-protein scores were associated with faster rates of cognitive and functional decline as assessed by the Clinical Dementia Rating (CDR) Sum of Boxes (higher scores ∼ clinical progression) over a median of 2.4 years (25^th^ − 75^th^ percentile: 1.9-5.1 years; PC1: standardized b=-0.13, p=0.0005; PC2: standardized b=-0.20, p<0.0001; PC3: standardized b=-0.08, p=0.02; **Figures 5D-F**). Moreover, lower multi-protein scores were associated with faster rates of grey matter atrophy and cognitive decline, as assessed by the Montreal Cognitive Assessment (MoCA) and episodic memory and executive function composite scores (**Supplemental Table 19**). Results were robust to adjustment for age, sex, cohort, race/ethnicity, *APOE*-ε4 status, smoking status, BMI, and area deprivation index (**Supplemental Table 19**).

### Multi-protein scores of neurostructural phenotypes quantified in midlife are associated with subsequent cognitive impairment and AD biomarkers up to two decades later

Changes in candidate AD biomarkers linked to cognitive endpoints are more likely to represent biological drivers of AD. To evaluate dynamics of the 3 multi-protein scores, we turned to CARDIA, a 40-year longitudinal cohort study with serial circulating proteomics (Olink) over 15 years (Exam Year 15 to Year 30; mean age 40 to 55 years; see **Methods**). In addition to serial proteomic profiling, established plasma biomarkers of AD (pTau-217, Aβ42 and Aβ40) were quantified at Year 35^33^ (mean age 60 years) with the following proportions of negative, intermediate, and positive results, respectively (defined in **Methods**): pTau-217 81%, 15%, 3.8%; pTau-217/ Aβ42 ratio 85%, 8.1%, 6.8%; Aβ42/ Aβ40 ratio 18%, 69%, 13%. The low proportion of pTau-217 positivity is consistent with the relatively young age of the cohort and the expected low prevalence of overt AD pathology.

Consistent with the *cross-sectional* inverse association with age in UK Biobank, we observed a general decrease in multi-protein scores *longitudinally* over time in CARDIA (**Figure 5G**). While scores at Year 15 (mean age 40) were not significantly associated with neurostructural traits at Year 30 (mean age 55), a higher multi-protein score of cerebrovascular health (PC3) at Year 15 was associated with higher cognitive performance ≈15 years later (**Figure 5H**, **Supplemental Table 20**). On average, individuals with <u>higher</u> multi-protein scores of white matter integrity (PC2) in midlife (Year 15) had <u>lower</u> pTau-217 or pTau-217 to Aβ42 ratio ≈2 decades later (Year 35, mean age 60). Importantly, multi-protein scores <u>across midlife</u> (approximately ages 40-55 years) showed substantially greater inter-individual variability than conventional AD biomarkers measured at Year 35 (**Supplemental Figure 4**), indicating that multi-protein scores may capture biologically informative differences among individuals during a period when AD biomarker abnormalities remain relatively uncommon. Demographic (age, sex, race) and metabolic (BMI, A1c) factors accounted for the largest share of explained variance in multi-protein scores (**Supplemental Figure 5**), consistent with neurostructural variation reflecting a cumulative influence of diverse metabolic and systemic processes on brain integrity^34,35^. Of note, most variance in the multi-protein scores remained unexplained by measured demographic, clinical, metabolic, or biomarker factors (**Supplemental Figure 5**), suggesting that the multi-protein scores capture additional biological variation not fully represented by conventional risk markers.

### Dynamicity in multi-protein scores of neurostructural phenotypes and AD phenotypes and risk

Nearly two decades of longitudinal follow-up in CARDIA allowed us to examine associations of longitudinal changes in multi-protein scores over time with cognitive and structural phenotypes. While the multi-protein scores at Year 15 correlated with later years, the correlations weakened with increasing time intervals (**Supplemental Figure 6**). In longitudinal models, both *higher* multi-protein scores at Year 15 (mean age 40) <u>and</u> a *flatter trajectory* over the subsequent 3 measurements spanning 15 years (more positive participant-specific slope = less decline) were associated with higher cognitive performance, higher grey matter volume, and a more favorable AD biomarker profile (lower pTau-217; **Figure 5H**; **Supplemental Figure 7**; **Supplemental Table 21**). Despite temporal sequence of measures (proteomics, Years 15-30; cognitive, imaging at Year 30; AD biomarkers, Year 35), participant-specific slopes of multi-protein scores from Year 15-to-30 exhibited consistent associations with cognitive and neurostructural phenotypes. In general, adjustment for multi-protein scores at Year 15 attenuated the relations of Year 35 AD biomarkers with cognitive and neurostructural phenotypes in CARDIA (**Supplemental Figure 8, Supplemental Table 22**). There was no statistical evidence for interaction between Year 35 AD biomarkers and Year 15 multi-protein scores or participant-specific slopes on cognitive performance. In contrast to findings from younger cohorts linking multi-protein scores to cognitive decline (VMAP, CARDIA), AD biomarkers (CARDIA), and incident dementia (UK Biobank), multi-protein scores in the Knight ADRC cohort (mean age 74 years; median follow-up 4 years, 1,009 participants with 191 AD cases) were not associated with incident AD (**Supplemental Figure 9**, **Supplemental Table 23**).

### Single-nucleus RNA sequencing (snRNA-seq) in human brain across individuals with and without AD provides brain- and cell type-specific context for the neurostructural proteome

Single-nucleus RNA-seq pre-processing and profiling were performed on superior frontal gyrus samples from 101 donors (66 with AD). Participants were volunteers enrolled in the Arizona Study of Aging and Neurodegenerative Disorders (AZSAND), a longitudinal study that has been conducted since 1996. After quality control to filter out low-quality nuclei and doublets, 481,840 nuclei were kept for subsequent gene expression analyses and genetics-anchored transcriptome-wide association studies (TWAS; **Methods**). Unsupervised clustering identified several cell clusters (**Figure 8A**), which underwent annotation using a reference snRNA-seq panel (syn18485175), including microglia, astrocytes, excitatory neurons, inhibitory neurons, oligodendrocytes, endothelial cells, pericytes, and oligodendrocyte progenitor cells (OPCs). The “grey matter structure” (PC1) associated proteins showed enrichment across multiple major cell subpopulations, including microglia, astrocytes, endothelial cells, and OPCs (**Figures 9A-B**), revealing a broad cellular signature consistent with the complex cellular organization of the affected structure. The “white matter integrity” PC2-associated proteins displayed a more concentrated cellular architecture, characterized by predominant enrichment in astrocytes (**Figures 9C-D**), whereas the “cerebrovascular health” PC3-associated proteins were preferentially enriched in OPCs (**Figures 9E-F**). Collectively, these enrichment patterns suggest that the major neurostructural PCs are associated with differential cellular contributions rather than *discrete* cell-type specificity. PC1 reflected a broadly distributed multicellular signature whereas PC2 and PC3 showed comparatively stronger contributions from astrocyte-related and OPC-related biology, respectively, highlighting distinct biological axes of neurostructural variation.

**Figure 8.**
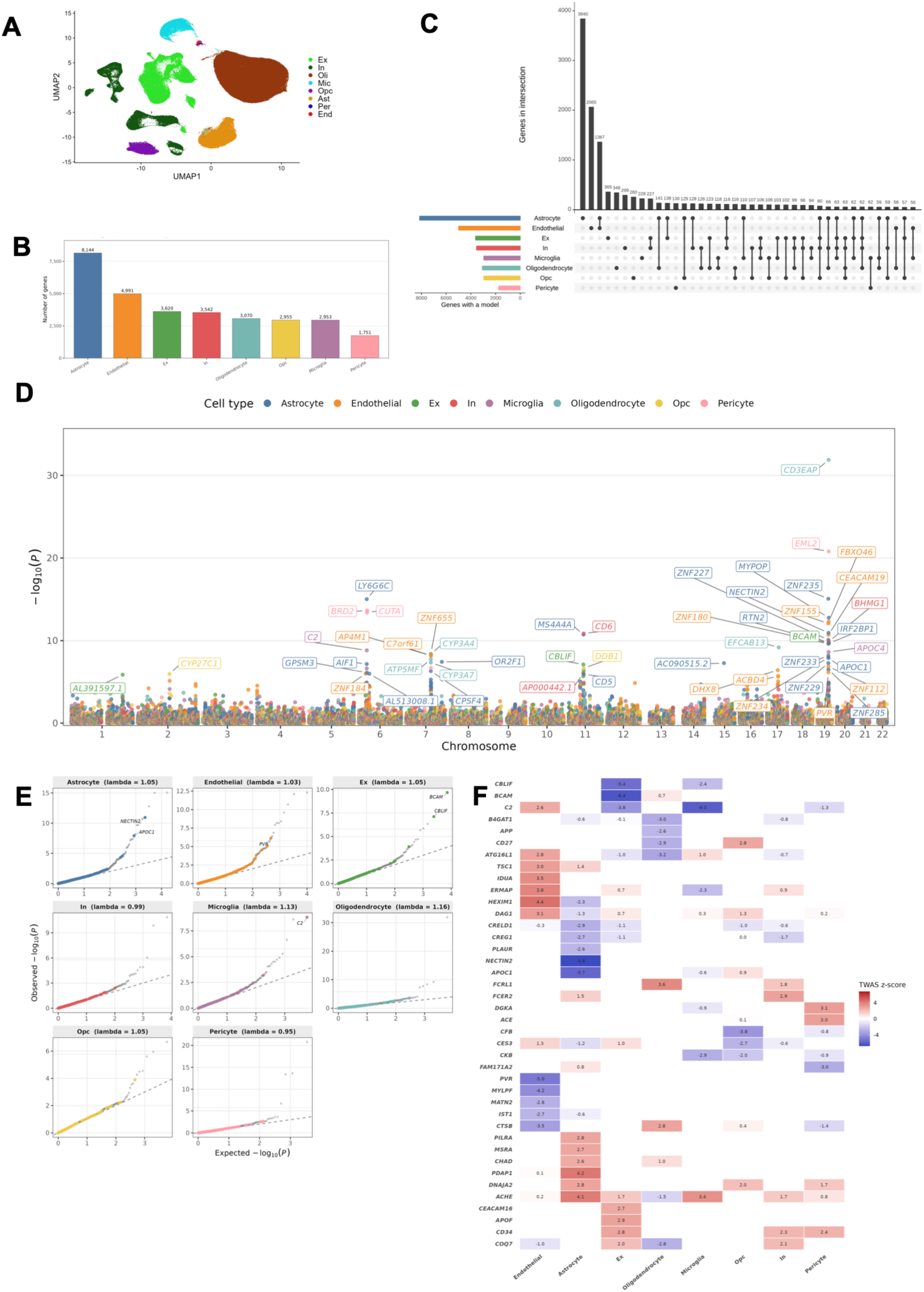
Cell specific genetic associations with Alzheimer’s disease. **(A)** Unsupervised clustering identified several clusters among the cell subpopulations. The clusters were annotated using a reference snRNA-seq panel (syn18485175). Cell clusters include Mic: microglia, Ast: astrocytes, Ex: excitatory neurons, In: inhibitory neurons, Oli: oligodendrocytes, End: endothelial cells, Per: pericytes, and Opc: oligodendrocyte progenitor cells. **(B)** Genetically-anchored single-nucleus transcriptome-wide association study models of gene expression were trained using the cis-regulatory region of a gene for 8144 genes in astrocytes, 4991 in endothelial cells, 3620 in excitatory neurons, 3542 in inhibitory neurons, 3070 in oligodendrocytes, and 2953 in microglia. **(C)** The global pattern of resTWAS gene coverage across cell types displayed notably high cell type specificity, indicating strong context dependence of genetic regulation of gene expression. Astrocytes displayed the highest level of gene model sharing with other cell types, consistent with the cell type’s role in regulating neuronal, vascular, metabolic, and inflammatory processes. **(D)** We conducted resTWAS of AD using a large-scale GWAS, identifying significant cell type anchored gene-level associations across the major cell subpopulations in the brain. *CD3EAP*, a gene located on chromosome 19, in oligodendrocytes showed the most significant resTWAS association with AD (p<10^−30^). **(E)** The resTWAS of AD showed cell type dependent excess for association signals with AD, as visualized by leftward shift in the Q-Q plot, with our neurostructural-associated proteins among the most significant associations (Bonferroni-adjusted p<0.05) in astrocytes (*NECTIN2*, *APOC1*), endothelial cells (*PVR*), excitatory neurons (*BCAM*, *CBLIF*), and microglia (*C2*) for the transcripts encoding the proteins. **(F)** The genes encoding the proteins displayed varying levels of cell type specificity of resTWAS associations. In general, the effect sizes on AD displayed high concordance across the multiple cell subpopulations in which the gene is genetically regulated.

**Figure 9.**
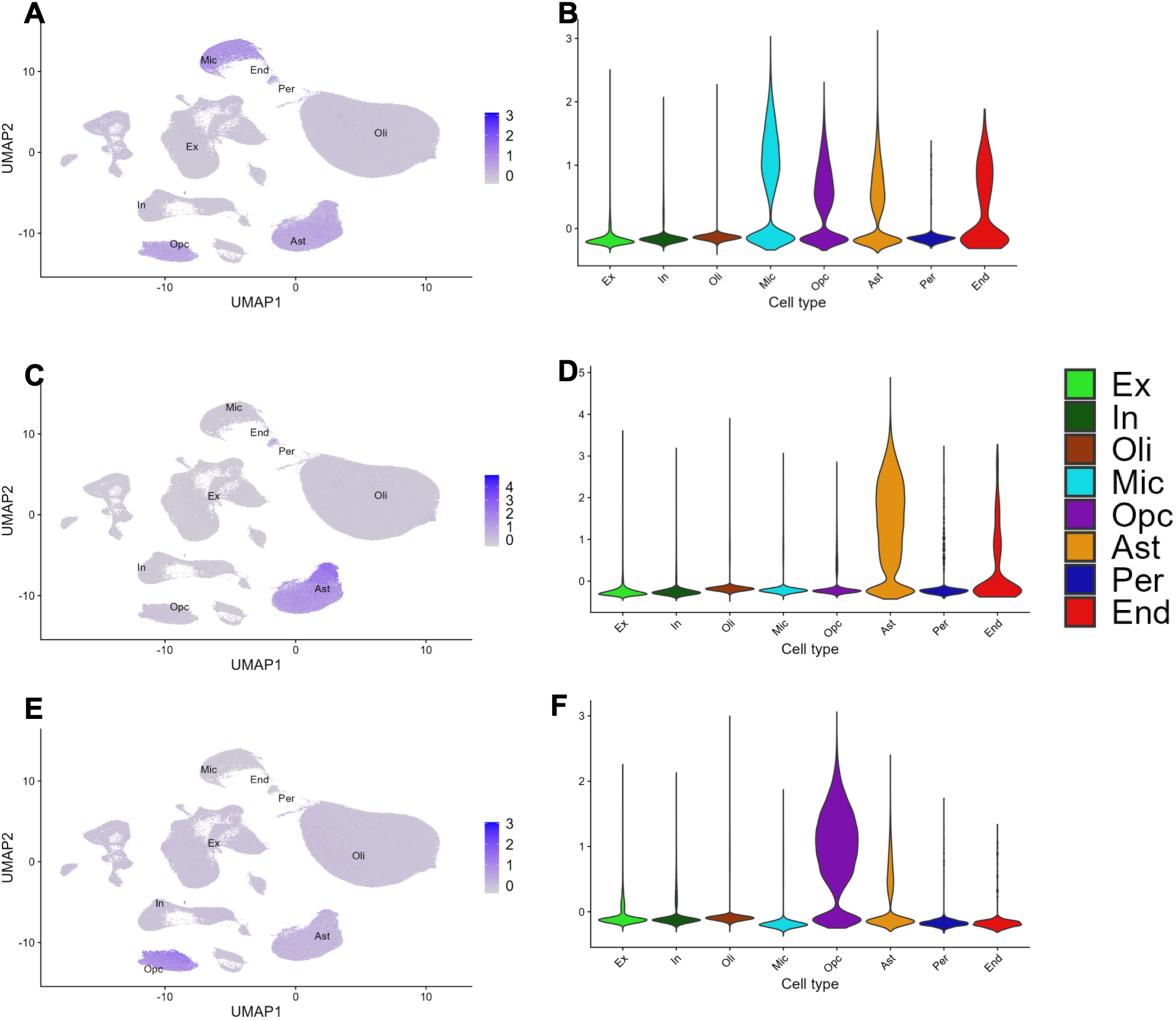
The neurostructural proteome is enriched by cell type. **(A, B)** The grey matter structure (PC) associated proteins showed enrichment across multiple major cell subpopulations, including microglia, astrocytes, endothelial cells, and oligodendrocyte progenitor cells. **(C, D)** White matter integrity (PC2) associated proteins displayed a more cell type specific pattern, with predominant enrichment in astrocytes. **(E, F)** Cerebrovascular health (PC3) associated proteins were preferentially enriched in oligodendrocyte progenitor cells. Cell clusters include Mic: microglia, Ast: astrocytes, Ex: excitatory neurons, In: inhibitory neurons, Oli: oligodendrocytes, End: endothelial cells, Per: pericytes, and Opc: oligodendrocyte progenitor cells. Submitted Manuscript: Confidential

### Single-nucleus transcriptome-wide association study

We used the snRNA-seq data paired with whole-genome sequencing to train genetic models of gene expression at the level of individual cell types. By applying a genetically-anchored single-nucleus resolution-enhanced TWAS (resTWAS) approach to GWAS summary statistics (**Methods**), we can identify genes, including those encoding proteins within the neurostructural proteome, whose genetically regulated expression is associated with genetic susceptibility to AD, as well as the specific cell types in which these genes may exert their effects. Genetic models of gene expression were developed using the *cis*-regulatory region (1 Mb) of a gene for 8,144 genes in astrocytes, 4,991 in endothelial cells, 3,620 in excitatory neurons, 3,542 in inhibitory neurons, 3,070 in oligodendrocytes, and 2,953 in microglia, demonstrating a substantial number of genetically-predicted molecular traits in each cell subpopulation (see **Figure 8B** for gene model counts across all cell types). The median number of SNP features for optimal prediction was 11 across the major cell subpopulations except for the excitatory and inhibitory neurons which had a median of 12 (**Supplemental Figure 10A**), indicating substantial allelic heterogeneity, with a large fraction of gene expression traits across cell types optimally predicted by >50 SNPs (**Supplemental Figure 10B**). Nearly 49% of the genes (*n* = 7,535) had genetic models in a single cell type, and 46% could be modeled genetically in 2 to 4 cell types (**Figure 10**). The overall pattern of resTWAS gene coverage across cell types showed high cell type specificity (**Figure 8C**), indicating strong context dependence of the genetic regulation of transcript levels. Astrocytes displayed the highest level of gene model sharing with other cell types, consistent with the cell type’s broad involvement in regulating neuronal, vascular, metabolic, and inflammatory processes. Astrocytes and endothelial cells showed the highest number (1,387) of overlapping resTWAS gene models, in accordance with their joint roles in blood-brain-barrier integrity, vascular regulation, and neurovascular coupling.^36^

**Figure 10.**
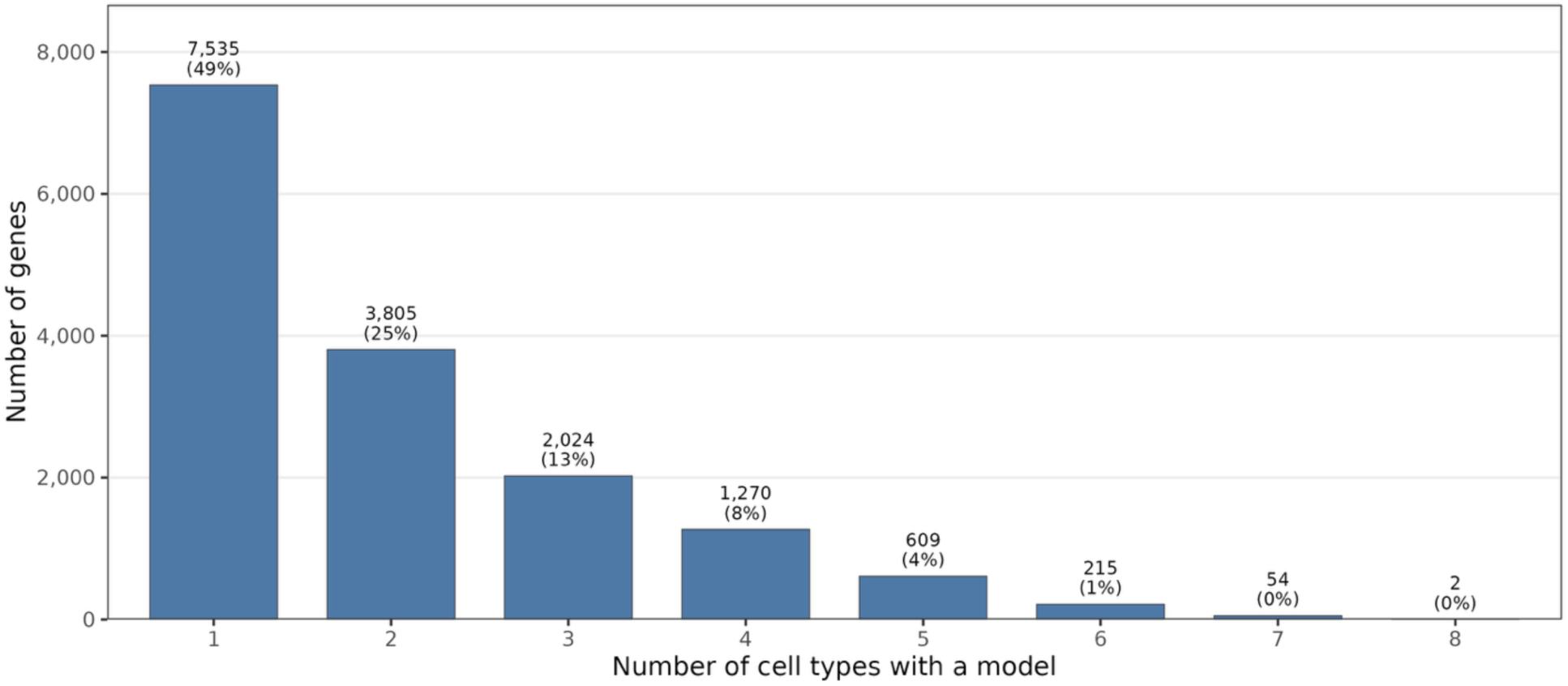
Genetic models per cell type. Approximately 49% of the genes (*n* = 7535) had genetic models in only one cell type, and 46% could be modeled genetically in 2 to 4 cell types in decreasing numbers, indicating cell type specificity of the genetically determined component.

### resTWAS of AD implicates the neurostructural proteome in cell type-specific AD genetic risk

We performed resTWAS using a large-scale GWAS of AD (111,326 clinically diagnosed/‘proxy’ AD cases and 677,663 controls)^37^, identifying significant cell type anchored gene-level associations across the major cell subpopulations in the brain (**Figure 8D; Supplemental Table 24; Methods**). *CD3EAP*, a gene located on chromosome 19, in oligodendrocytes showed the most significant resTWAS association with AD (P<10^−30^). While the *APOE* gene has historically been the subject of considerable research, the *CD3EAP* locus (rs2229918) has been shown to be among the most powerful genetic risk factors, along with *APOE*, for AD risk and progression^38^.

The resTWAS showed cell type-dependent enrichment for association signals with AD, with our neurostructural proteome among the most significant associations (Bonferroni-adjusted p<0.05) in astrocytes (*NECTIN2*, *APOC1*), endothelial cells (*PVR*), excitatory neurons (*BCAM*, *CBLIF*), and microglia (*C2*) for the transcripts encoding the proteins (**Figure 8E** and **Supplemental Figure 11**). The genetically determined expression of *NECTIN2*, a gene with variants not in linkage disequilibrium with (i.e., independent of) *APOE*-ε4^39^ and previously reported to mediate hippocampal atrophy in AD^39^, was the most significant AD association (P=1.47×10^−11^), specifically in astrocytes, among the neurostructural proteomics-prioritized genes. *PVR*, a cell adhesion molecule that is the major DNAM-1 ligand on primary vascular endothelial cells^40^ and regulatory for transendothelial migration of immune cells,^41^ was the most significant AD association in brain endothelial cells (P=7.13×10^−7^) among the neurostructural proteome prioritized genes. The *NECTIN2* and *PVR* resTWAS models were specific to a single cell type (astrocytes and endothelial cells, respectively), indicating that the genetic regulatory effects were identified exclusively within particular cell subpopulations (and not in others). *CBLIF* (P=7.7×10^−8^ in excitatory neurons), a protein known for its critical role in vitamin B12 transport, may impact AD susceptibility by defending against excitotoxicity^42,43^ and blocking the accumulation of neurotoxic metabolites in vulnerable excitatory neurons.^44^

The full neurostructural proteome showed significant cell type-dependent enrichment for resTWAS associations with AD across the major cell subpopulations in the brain, except for inhibitory neurons (**Supplemental Figure 11; Methods**). Importantly, enrichment for AD-associated resTWAS signals within the neurostructural proteome remained robust after exclusion of chromosome 19 genes, including the APOE region, indicating that convergence with AD genetic architecture is not driven by the extended APOE region and extends beyond the effects of the strongest known AD susceptibility locus (**Supplemental Figure 12**). The genes encoding the neurostructural proteome displayed varying levels of cell type specificity of resTWAS associations, but in general, the effect sizes on AD displayed high concordance across the multiple cell subpopulations in which the gene is genetically regulated (**Figure 8F**). Finally, we observed that the pattern of substantial enrichment for gene-level associations in the major cell types among the neurostructural proteome was highly specific to AD, as no enrichment—and indeed, some depletion—was observed for another neurodegenerative disorder, Parkinson’s disease (PD) (**Figure 11; Methods**). Notably, AD-associated resTWAS signals identified within the neurostructural proteome showed no overlap with corresponding PD associations, supporting preferential convergence with AD-related genetic architecture rather than nonspecific neurodegenerative or brain-aging processes.

**Figure 11.**
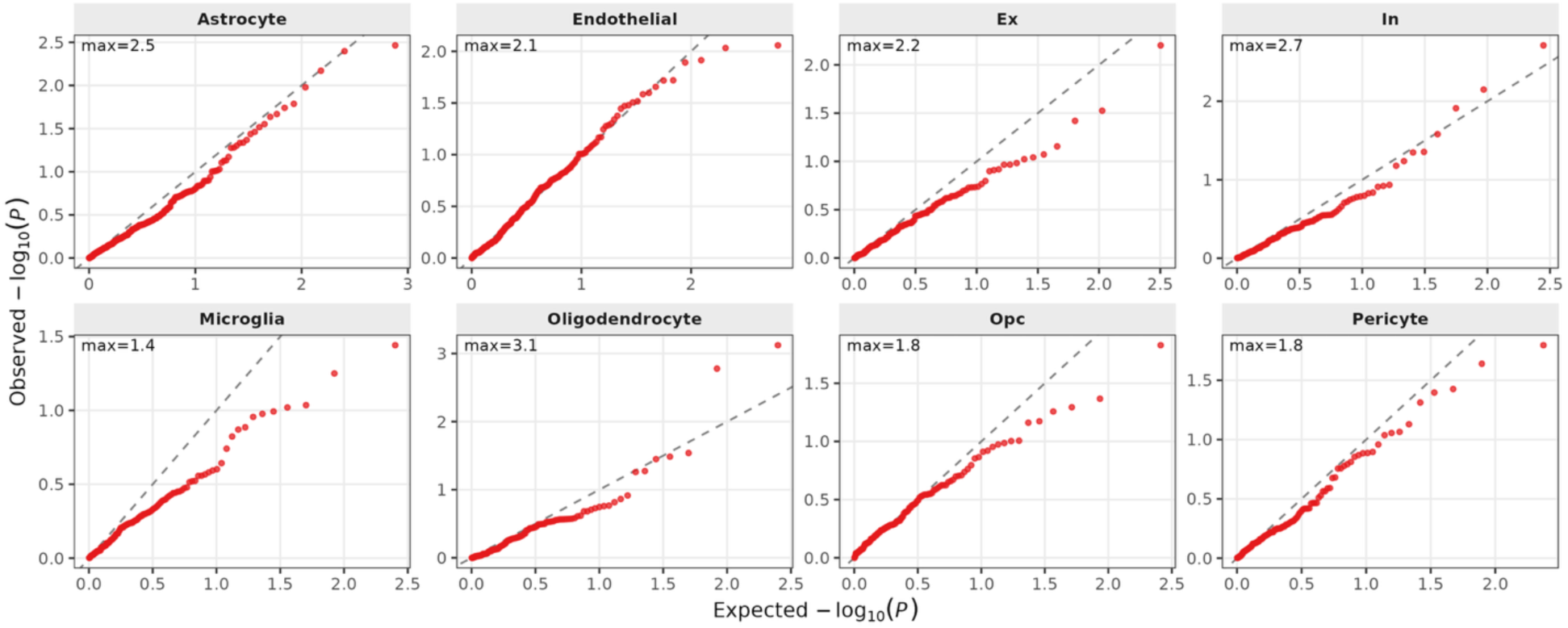
The neurostructural proteome is not enriched in Parkinson’s disease. The resTWAS of PD did not show cell type dependent excess for association signals with PD.

## DISCUSSION

Defining and targeting early pathobiology is critical to intercepting AD before the onset of progressive cognitive decline. Despite the high positive predictive value of circulating neuropathologic biomarkers for AD, sensitivity for early AD detection may be limited^8^. Here, we integrated MRI phenotypes, proteomics, longitudinal cohort epidemiology, and single-nucleus brain genomics to identify a neurostructural proteome that converges on AD-related genetic, cellular, and biomarker signatures and that identifies a midlife molecular state that anticipates future AD susceptibility. The neurostructural proteome reflected AD pathobiology, converged on AD-specific genetic liability at single-nucleus resolution, and multi-protein scores of neurostructural phenotypes were strongly associated with incident AD independent of polygenic risk. The neurostructural proteome featured both metabolic-inflammatory and neurophysiological mechanisms, with enrichment in both brain and peripheral metabolic tissues (blood, liver) and genetic associations with key AD-related phenotypes. Multi-protein scores measured *near age 40* (well before typical pTau-217 assessment) were associated with cognitive performance and AD biomarker profiles 15 years later. Finally, using single-nucleus transcriptomics and genomics, we demonstrated preferential enrichment in astrocyte, microglia, and endothelium cells among the genes encoding for the circulating neurostructural proteome through resTWAS. AD-associated genetically determined expression signals identified through resTWAS within the neurostructural proteome comprised a mixture of secreted-immune metabolic mediators (e.g., *APOC1*^45^, *C2*^46^), endothelial/cell-adhesion molecules (e.g., *PVR*^40^), and intracellular regulators (e.g., *PDAP1*), implicating coordinated immune, metabolic, and neurovascular processes in AD. Importantly, resTWAS implicated the neurostructural proteome in AD but <u>not</u> PD loci from GWAS. AD-associated signals identified with the neurostructural proteome showed no overlap with those observed for PD, supporting preferential convergence with AD genetic architecture rather than shared neurodegenerative processes. Notably, AD-associated resTWAS enrichment remained robust after exclusion of chromosome 19 genes, indicating that the genetic support for AD within the neurostructural proteome was not solely driven by the extended APOE region. Collectively, these findings suggest that the circulating neurostructural proteome captures midlife biological processes preferentially associated with future AD susceptibility, detectable decades before conventional biomarkers are typically elevated.

Classically, the earliest lesion in AD is thought to involve Aβ processing (at a cognitively preserved phase), followed by metabolic imaging abnormalities, neurodegeneration, and tau accumulation^47^. Major advances in AD leverage this paradigm, including deploying amyloid-modifying therapies at diagnosis and using circulating pTau-217 (and newer microtubule-binding region MTBR-tau^48^) for diagnosis. Nevertheless, diagnostic use of AD biomarkers in individuals without cognitive impairment is not currently implemented. Furthermore, in a recent study of 2,148 individuals free of dementia age 60 and older, Grande and colleagues demonstrated high negative but low positive predictive values for tau- and amyloid-based biomarkers can be augmented by addition of select proteins^8^. Our approach integrated neuroimaging, proteomics, and life-course cohorts (some decades before classic pTau-217 positivity) to suggest this adjunctive paradigm for AD detection in a younger at-risk population. Brain imaging is an integral part of AD diagnosis^49^ and molecular discovery^50–53^. Neurostructural endophenotypes are optimal to capture subtle, upstream alterations relevant to AD before its clinical manifestations. Indeed, prior AD proteomic discovery efforts anchored to MRI phenotypes^54–57^ or diagnosis have some consistency with our findings (e.g., GFAP, NEFL, GDF15, CRLF1, ADAMTSL2 predictive of AD years before clinical onset^58,59^). Nevertheless, most large longitudinal proteomic studies of sporadic AD generally start at an older age than CARDIA (mean age ≈55-60 years or older at time of proteomics or imaging^54,59,60^). The epidemiology of AD highlights the importance of midlife (ages ≈40-55 years; well before the typical detection of pTau or Aβ^33^) to later cognitive impairment^11^. Furthermore, multi-protein scores of neurostructural phenotypes developed in UK Biobank, when measured at a mean age ≈40 years in CARDIA, were associated with AD biomarkers and cognitive performance nearly two decades later. This finding highlights biological variation associated with subsequent AD-related outcomes that is detectable years before conventional biomarker positivity.

A primary question in organ-specific *circulating* biomarker development is whether identified biomarkers reflect organ-specific dysfunction or are an “epiphenomenon” of related disorders (like aging, obesity, etc.). Proteomic studies have recently addressed this query by labeling individual proteins as organ-specific via their tissue-specific transcriptional profiles^61^. Our results suggest both peripheral and neurologic origins for identified proteins. As part of this work, we developed a single-nucleus genomics atlas of brain transcription, demonstrating candidates within the neurostructural proteome enriched at the transcriptional level in astrocytes, microglia, and endothelial cells—brain cell types central to AD. Furthermore, single-nucleus TWAS models mapped the neurostructural proteome to cell type-specific genetic liability to AD (but not PD). Moreover, the blood and liver also served as an enriched source of the neurostructural proteome, consistent with studies demonstrating both peripheral^62,63^ and brain metabolic abnormalities^64^ in AD pathogenesis. A substantial fraction of the explainable variance in our multi-protein scores was attributable to demographic and metabolic factors, with proteins in signatures also specifying metabolic-inflammatory pathways (AMY2B, TREH, LIPF, IL6, IL17C, CCL4, CXCL9, AGER). The presence of metabolic-inflammatory proteins in molecular signatures of AD is analogous to other chronic disorders^14^, where early metabolic modulation can interrupt accumulation of risk and clinical disease (e.g., PCSK9 inhibition in high cardiovascular risk^13^). Indeed, metabolic intervention (e.g., diet, exercise) even in the seventh decade of life improves cognitive and executive performance^65,66^, and midlife epidemiology has linked physical activity to lower risk of long-term dementia^67^. Furthermore, the finding that longitudinal changes in multi-protein scores are linked to AD-relevant phenotypes nearly two decades later suggests the potential for early disease modification. Collectively, these findings provide biological context to epidemiologic findings, implicating the brain and systemic metabolism in early phases of AD pathogenesis.

Identifying diverse origins for AD from circulation has implications on therapy and prevention. A landmark proposal by De Strooper and Karran in 2016 highlighted an early “biochemical phase” of AD upstream of amyloid and tau biology, characterized by abnormal proteostatic mechanisms of aging^68^. While this report sparked a renaissance in multi-omics evaluation of AD^69–75^, a full translation to identify this early biochemical phase postulated a decade ago—from brain architecture to the proteome, cellular states, genetics, and eventually clinical AD—has not been resolved. Both genetic findings and longitudinal cohort analyses spanning ages 40 to 75+ point to a role for interconnected metabolic and neurostructural pathways in AD pathobiology, well before neuropathologic changes classically manifest. Importantly, beyond discovery-specific effects related to differences in cohort composition, covariate adjustment, and follow-up duration, the lack of replication in the older Knight ADRC cohort suggests that multi-protein scores capturing midlife molecular states may reflect early susceptibility-related biology that attenuates with aging, whereas markers directly linked to neuropathological burden remain informative closer to the clinical expression of AD. Indeed, while AD tissue biomarkers and brain proteins (NEFL, GFAP) predict transitions from cognitive impairment to AD well (in older age), they may not be as effective in the long pre-clinical, biochemical phase^8,76^. In addition, multi-morbidity development may confound circulating proteome with age^77^. Finally, from a therapeutic perspective, current landmark disease-modifying therapies in AD focus on clearance of established amyloid plaques (lecanemab^78^, donanemab^79^). Although these therapies slow the trajectory of cognitive decline, they do not measurably reverse established neurodegeneration and may have treatment-related side effects (e.g., amyloid-related imaging abnormalities). These findings are analogous to biomarker and therapeutic discovery in other conditions like cardiovascular disease, where biomarkers of tissue injury or stress apparent <u>during a clinical presentation</u> (like cardiac troponin) may not serve as optimal biomarkers or therapeutic targets at the earliest interruptible phase.

Our study is strengthened by its translational approach spanning multi-modal data types, longitudinal epidemiology, and brain single-nucleus genomics to generate a biologically plausible markers of AD rooted in human neurostructural phenotypes. Nevertheless, several limitations warrant mention. While we did not have access to CSF or brain proteomics to measure the neurostructural proteome in the brain, single-nucleus genomics/transcriptomics implicated genes encoding the neurostructural proteome in genetic signals for AD (including brain-specific region TWAS), suggesting concordant pathways between brain and blood are relevant and represented in the periphery. In addition, CARDIA did not assess AD biomarkers at its earlier examinations (at the time of proteomics), although cognitive impairment or detectable pTau217 at age ≈40 is uncommon. Furthermore, while differences in proteomic profiling methods by cohort are noted (aptamer-vs. antibody-based), genetic replication of an identified neurostructural proteome in resTWAS studies alongside concordant results across similar platforms (in UK Biobank, CARDIA, VMAP) support biological importance. Since the neurostructural proteome was initially derived in the UK Biobank, replication in additional large-scale imaging-proteomic datasets will enhance the findings across diverse populations and study designs. Nevertheless, our principal biological conclusions were supported across multiple independent cohorts and associations were replicated using external cohorts with longitudinal cognitive trajectories, incident dementia, plasma AD biomarkers, and single-nucleus transcriptomic and genetic data. Finally, translation of these proteomic biomarkers to clinical relevance will require rapid absolute quantification and establishment of thresholds to guide AD risk assessment.

In summary, across thousands of individuals with MRI-derived traits, blood proteomics, longitudinal cohorts spanning two decades, single-nucleus brain genomics, and human genetics, we identified a circulating neurostructural proteome in midlife associated with cognition and AD-relevant endpoints decades later. Identified proteins span brain and genetic features strongly linked to AD biology in large human studies and tissue-based single-nucleus transcriptomics. Importantly, multi-protein scores were associated with subsequent cognitive impairment over both short- and long-term time horizons. Collectively, these results define a circulating neurostructural proteome that links midlife molecular variation to later-life cognitive decline, AD-related biomarker status, and AD genetic architecture, expanding opportunities to investigate disease-relevant processes decades before clinical manifestation.

## Acknowledgments

We are grateful to the Banner Sun Health Research Institute Brain and Body Donation Program of Sun City, Arizona for the provision of human biological materials. The Brain and Body Donation Program has been supported by the National Institute of Neurological Disorders and Stroke (U24 NS072026 National Brain and Tissue Resource for Parkinson’s Disease and Related Disorders), the National Institute on Aging (P30 AG19610 and P30AG072980, Arizona Alzheimer’s Disease Center), the Arizona Department of Health Services (contract 211002, Arizona Alzheimer’s Research Center), the Arizona Biomedical Research Commission (contracts 4001, 0011, 05-901 and 1001 to the Arizona Parkinson’s Disease Consortium) and the Michael J. Fox Foundation for Parkinson’s Research. Study data were obtained from the Vanderbilt Memory and Aging Project. Data were collected and processed by Vanderbilt Memory and Alzheimer’s Center investigators at Vanderbilt University Medical Center funded by the following grants: R01-AG034962, R01-AG056534, R01-NS100980, R01-AG062826, and Alzheimer’s Association IIRG-08-88733. The Coronary Artery Risk Development in Young Adults Study (CARDIA) is conducted and supported by the National Heart, Lung, and Blood Institute (NHLBI) in collaboration with the University of Alabama at Birmingham (75N92023D00002 & 75N92023D00005), Northwestern University (75N92023D00004), University of Minnesota (75N92023D00006), and Kaiser Foundation Research Institute (75N92023D00003). This manuscript has been reviewed by CARDIA for scientific content. This study was made possible by access to the UK Biobank resource (application ID 57492). This work was supported by access to equipment made possible by the Hope Center for Neurological Disorders, the NeuroGenomics and Informatics Center (NGI: https://neurogenomics.wustl.edu/) and the Departments of Neurology and Psychiatry at Washington University School of Medicine. These studies were supported by the Functional Multi-Omics for Diabetes Core of the Vanderbilt Diabetes Research and Training Center (P30DK020593 to ERG) We would like to thank all individuals for their participation. Microsoft Copilot was used for copy-editing. The authors are responsible for the manuscript and its contents.

## Funding

National Institutes of Health grant F30-AG085905 (HAA)

National Institutes of Health grant T32-GM007347 (HAA)

National Institutes of Health grant R01AG092663 (RVS, ERG)

National Institutes of Health grant P30DK020593 (RVS, ERG)

National Institutes of Health grant R01MH126459 (ERG)

National Institutes of Health grant U01DK140952 (ERG)

National Institutes of Health grant U24 NS072026 (AZSAND)

National Institutes of Health grant P30 AG19610 (AZSAND)

National Institutes of Health grant R01-AG034962 (ALJ)

National Institutes of Health grant R01-AG056534 (ALJ)

National Institutes of Health grant K24-AG046373 (ALJ)

National Institutes of Health grant UL1-TR000445 (VMAP)

National Institutes of Health grant UL1-TR002243 (VMAP)

National Institutes of Health grant P20-AG068082 (VMAP)

National Institutes of Health grant P30-AG086403 (VMAP)

National Institutes of Health grant R01-AG064614 (CC)

National Institutes of Health grant U01-AG084514 (CC)

National Institutes of Health grant P01-NS131131 (CC)

National Institutes of Health grant R01-AG078964 (CC)

National Institutes of Health grant R01-AG058501 (CC)

National Institutes of Health grant R01-AG071706 (CC)

National Institutes of Health grant P30-AG066444 (CC)

National Institutes of Health grant R01-AG064877 (CC)

National Institutes of Health grant P30AG066444 (Knight ADRC)

National Institutes of Health grant P01AG03991 (Knight ADRC)

National Institutes of Health grant P01AG026276 (Knight ADRC)

National Brain and Tissue Resource for Parkinson’s Disease and Related Disorders (AZSAND)

Arizona Alzheimer’s Disease Core Center (AZSAND)

Arizona Department of Health Services (AZSAND)

Arizona Alzheimer’s Consortium (AZSAND)

Arizona Biomedical Research Commission (AZSAND)

Arizona Parkinson’s Disease Consortium (AZSAND)

Michael J. Fox Foundation for Parkinson’s Research (AZSAND)

Scott Hamilton CARES Foundation (ERG)

Alzheimer’s Association IIRG-08-88733 (ALJ)

Richard Eugene Hickman Alzheimer’s Disease Research Endowment (VMAP)

Vanderbilt Memory and Alzheimer’s Center (VMAP)

Cure Alzheimer’s Fund (Knight ADRC)

## Author contributions

Conceptualization: HAA, TJH, ALJ, RVS, ERG

Methodology: HAA, ASP, XB, KT, QSW, TJH, ALJ, KVKJ, RVS, ERG

Investigation: ASP, HAA, XB, DA, AT, PL, EFE, BLP, SZ, QS, SM, EH, PZ, MWA, R Sale, YC, LC, AB, KT, AR, DL, KRP, NS, CB, LTD, BAL, CC, R Saloner, KY, TJH, ALJ, KVKJ, RVS, ERG

Visualization: HAA, ASP, XB, AT, PL, QS, PZ, MWA, RVS, ERG

Funding acquisition: HAA, CC, ALJ, ERG

Project administration: ASP, HAA

Supervision: KVKJ, RVS, ERG

Writing – original draft: HAA, ASP, RVS, ERG

Writing – review & editing: HAA, ASP, XB, DA, AT, PL, EFE, BLP, SZ, QS, SM, EH, PZ, MWA, R Sale, YC, LC, AB, KT, AR, JEF, MN, SD, DL, KRP, NS, CB, LTD, BAL, LH, CC, R Saloner, KY, QSW, TJH, ALJ, KVKJ, RVS, ERG

## Competing interests

A.S.P. is a co-inventor on patents or disclosures on molecular biomarkers of fitness, lung disease, dementia, cardiovascular diseases and phenotypes, and metabolic health. T.J.H. is a scientific advisor for Circular Genomics, Deputy Editor for Alzheimer’s & Dementia: Translational Research and Clinical Intervention, Section Editor for Alzheimer’s & Dementia, and co-founder of Precision-AD. A.L.J. is an advisor for Medtronic, and Chair of the Observational Study Monitoring Board for the Diverse-VCID: White Matter Lesion Etiology of Dementia in Diverse Populations Study. R.V.S. is supported by grants from the National Institutes of Health. R.V.S. has equity ownership in, is consultant for, and has support for travel from Thryv Therapeutics. He has research and travel support from Kardigan. He has research support from Bayer. R.V.S. is a co-inventor on patents or disclosures on molecular biomarkers of fitness, lung disease, dementia, cardiovascular diseases and phenotypes, and metabolic health, as well as use of RNAs (including spatial) as therapeutics and diagnostic biomarkers in disease and methods in metabolomics. E.R.G. has served as a consultant for Thryv Therapeutics and is a co-inventor on disclosures and patents for molecular signatures of cardiovascular and neurological phenotypes and metabolic health, use of RNAs as therapeutics and biomarkers in disease, and methods in metabolomics.

## Data, code, and materials availability

Data is available through correspondence with the respective sources, pending appropriate data use agreements and IRBs and subject to cohort-specific data sharing policies: (1) UK Biobank (publicly available at https://www.ukbiobank.ac.uk); (2) Knight ADRC (correspondence with CC); (3) GNPC (publicly available at https://discover.alzheimersdata.org/catalogue/datasets/e2f3536b-d97b-4303-89bd-6864200807a4); (4) VMAP data (correspondence with ALJ); (5) CARDIA data (through correspondence with CARDIA Coordinating Center at https://sites.uab.edu/cardia/); (6) AZSAND data (correspondence with KVKJ). No new materials or code were created for this study.

## Supplementary Materials

Materials and Methods

Figs. S1 to S12

Tables S1 to S24

References (80–119)

## Materials and Methods

This study utilizes data from several cohorts. Detailed reports of study recruitment, clinical assessment, and proteomic quantification have been published previously; key aspects relevant to this analysis are summarized here, with attribution to these prior publications given by this statement.^33,80–85^

### Study cohorts

#### UK Biobank (Discovery Cohort)

The UKB is a prospective cohort study of community-dwelling adults across the United Kingdom ranging from 40 to 69 years old at recruitment. For this project, we identified participants with Olink Explore 3072 proteomics data available (n=52,995). We created two cohorts for this project (with some participants overlapping both cohorts), including (1) 8,711 participants with complete cases neuroimaging data available for models relating proteomic measures to neurostructural phenotypes and (2) 9,973 participants with complete proteomics data for the 3 protein scores and follow-up data available for models leveraging proteomic risk scores to predict future dementia diagnosis. Within the UKB cohort employed for models relating proteomic measures to neurostructural phenotypes, participants ranged from 40 to 70 years old (54±8 years), 54% were female, 96% self-identified as White, 27% were carriers of the *APOE*-e4 allele, 40% identified as current or prior smokers, and 47% reported having an education level equivalent to a college degree or higher. Townsend deprivation index ranged from −6.3 to 9.1 (−1.7±2.8) and body mass index (BMI) ranged from 15.7 to 55.5 (26.5±4.3). At baseline, only 2 (<0.01%) participants included in models relating circulating proteins to neurostructural phenotypes had a diagnosis of all-cause dementia. For survival models (n=9,973), all participants with prevalent diagnoses of dementia were excluded, and overall participant characteristics were similar to the cohort described above (see **Supplemental Table 1** for full participant characteristics for all cohorts).

#### Knight Alzheimer’s Disease Research Center (Knight ADRC)

Data from Knight ADRC participants was used to replicate UKB Cox model results. Knight ADRC is a longitudinal cohort study of memory and aging that began in 1979.^82^ Participants are seen annually for examination and cognitive testing. For this study, we only included participants with proteomic data who were cognitively normal at baseline and individuals whose final diagnosis was either cognitively normal or Alzheimer’s disease (n=1,009, 74±10 years, 59% female). Since the Knight ADRC is one of the cohorts contributing to GNPC, differential abundance analyses were not performed using this dataset.

#### Global Neurodegeneration Proteomics Consortium (GNPC)

GNPC is the world’s largest collection of biofluid proteomics in participants with neurodegenerative diseases and normal cognition.^80^ For the current project, the harmonized GNPC dataset with SomaScan plasma proteomic data was leveraged (n=18,856). To reduce diagnostic ambiguity, as previously done,^80,86^ the analytic sample was restricted to participants with non-overlapping clinical diagnoses. This resulted in the exclusion of participants with multiple neurodegenerative diagnoses (n=150) as well as those with both MCI and AD (n=266). We defined participants as being cognitively normal if they were free from all labeled neurodegenerative conditions, free from diagnosed MCI, and were either explicitly labeled as normal in their respective cohorts and/or had a CDR of 0 (n=8,419 participants with no diagnosis who did not meet the criteria for cognitively normal were removed). We excluded participants with missing age (n=49). This resulted in a total analytic sample of 9,972 GNPC participants for differential abundance analyses (70±12 years, 56% female), including 2,890 AD, 613 PD, 214 FTD, 443 ALS, and 5,812 cognitively normal participants. A subset of 2,835 GNPC participants had non-missing values on all proteins required to calculate the multi-protein scores.

#### Vanderbilt Memory & Aging Project (VMAP)

VMAP is a longitudinal observational study of older adults based in Nashville, Tennessee ranging from 50 to 92 years old at study entry with up to 11-years of follow-up data available.^87^ VMAP participants included in the current project were leveraged from both the Legacy Cohort (recruitment 2012-2014, up to 12-years follow-up data available) and Expansion Cohort (recruitment 2021-present, up to 3-years follow-up data available). VMAP cohort-specific inclusion and exclusion criteria have been previously described in detail.^83,87,88^ For the current project, a total of 556 VMAP participants were included (66±9 years, 53% female), including 62 with MCI (CDR=0 or 0.5 and impairment on neuropsychological assessment),^89^ 10 with early MCI (CDR=0.5 without impairment on neuropsychological assessment),^90^ and 484 who were cognitively unimpaired at baseline (CDR=0 and no impairment on neuropsychological assessment). Of the 556 VMAP participants included in models, 370 had longitudinal follow-up data available (i.e., two or more distinct timepoints for cognitive and neurostructural outcomes).

#### Coronary Artery Risk Development in Young Adults (CARDIA)

CARDIA is a population-based, longitudinal prospective cohort study with aims to investigate the development of cardiovascular disease risk factors from young adulthood to midlife.^85^ The study enrolled 5,115 participants at four field centers (Birmingham, AL; Chicago, IL; Minneapolis, MN; and Oakland, CA) in 1985-86, balancing recruitment by sex, age, race, and education level. Participants were contacted annually and serial in-person examinations were performed at 2-5 year intervals. For this study, we included 1,831 participants with plasma proteomics data available from Years 15, 20, 25, and 30 or had AD biomarkers quantified at Year 35. Cognitive testing and brain MRI were performed at Year 30 and AD biomarkers were quantified at Year 35.^33,91^ We used the following definitions for biomarker status: pTau-217/Aβ42 − negative (≤0.0055), intermediate (0.0056–0.0086), and positive (>0.0086); pTau-217 − negative (≤0.128 pg/mL), intermediate (0.129–0.300 pg/mL), and positive (>0.300 pg/mL); Aβ42/40 ratio − negative (≥0.1053), intermediate (0.0820–0.1052), and positive (<0.0820).^33^

#### Single-nucleus RNA-seq data

scRNA-seq and corresponding whole-genome sequencing data used in the preparation of this article were obtained from the Arizona Study of Aging and Neurodegenerative Disorders (AZSAND) database (www.brainandbodydonationprogram.org).

### Plasma proteomics

Plasma proteomic data from all cohorts, as described in detail below, were mean centered and standardized to unit variance prior to analysis.

UKB plasma proteomic data were generated using the Olink Explore 3072 platform, as previously described.^92^ The current study included plasma proteomic data captured at baseline (instance 0, years 2006-2010). Protein assays with >50% missing data were removed (n=3), and missing data was not imputed for the remaining assays.

Knight ADRC plasma proteomic data were generated from the SomaScan 7K platform. Protein measurements were log_10_-transformed. No outlier removal or imputation was performed. If multiple aptamers targeted the same protein, the aptamer with the highest Spearman correlation with Olink measurements was selected.^93^ No missing values were present in the proteomic matrix used for the final analytic cohort.

GNPC plasma proteomic data were generated using SomaScan 7K. Protein measurements were log_2_ transformed, and extreme outliers (i.e., >5 SD above or below the mean across the full dataset) were removed, as previously described.^80^ If multiple aptamers targeted the same protein, the aptamer with the highest Spearman correlation with Olink measurements was selected.^93^ For differential expression analysis at the individual protein level, to mitigate technical variability introduced by different proteomic platforms (i.e., Olink vs. SomaScan), proteins with Spearman’s correlation of <0.5 with Olink 3072 measurements were excluded.^93^

VMAP plasma proteomic data were generated using the Olink Explore 3072 platform, as previously described^83^. Within each of the three batches, proteins were removed if they did not have a ‘pass’ for quality control and assay warnings as provided by Olink. Following the removal of proteins based on Olink provided warnings, proteins with an inter-plate coefficient of variation >50%, or >50% missing data were removed from each batch. Aligning with how Olink calculates the percentage of missing data, datapoints were considered missing if no value was provided, or the value provided was below the limit of detection. Assuming a protein had no more than 50% of datapoints either not captured or below the limit of detection, datapoints that were below the limit of detection remained as is and were not imputed or removed. A final within-batch filter was applied, removing samples with >10% missing datapoints. For proteins measured across multiple panels, a mean value was calculated after performing within-batch quality control.

CARDIA proteomic data were generated using the Olink Explore HT platform. Quality control was performed separately within each of the eight Olink HT assay blocks. To remove inter-plate technical variation, NPX values were batch-corrected across plates using ComBat (*sva* package^94^), with plate as the batch variable and study year retained as a covariate in the model. Multivariate outliers were then identified within each block via PCA of the block’s protein matrix: a sample was flagged as an outlier if any of its first ten PC scores exceeded five standard deviations in absolute value. Flagged samples were set to missing for that block only. This was iterated until no further outliers were detected. We performed additional data fidelity and agreement check on the proteome sample identifiers from two separate data sources (Northwestern University, University of Alabama at Birmingham) and removed five samples with matching conflicts. Our final dataset post QC consisted of n=8,234 total proteome samples across 3,259 subjects (four CARDIA study timepoints). Following batch correction and outlier removal, each protein was standardized (z-scored) across all subjects and timepoints pooled together, so that every protein had mean zero and unit variance across the full dataset. The cleaned, normalized data were used to derive three protein scores (PC1 Grey Matter Structure, PC2 White Matter Integrity, PC3 Cerebrovascular Health) by projecting onto UKB-derived lasso weights. Of the weighted proteins contributing to each score, all 84 were available for PC1, 99 of 102 for PC2 (3 missing), and 132 of 137 for PC3 (5 missing).

### Brain magnetic resonance imaging

Analyses to derive a dementia proteomic risk score made use of AD-sensitive brain IDPs generated by an imaging-processing pipeline developed and run on behalf of UKB, as previously described.^95,96^ Briefly, ≈100,000 UKB participants were invited at instance 2 (beginning in 2014) to undergo scanning at one of three dedicated imaging centers on a Siemens Skyra 3T with a 32-channel receive head coil^95^. T1-weighted images were acquired using a 3D MPRAGE acquisition with the following parameters: spatial resolution=1×1×1 mm^3^, inversion time=880 ms, repetition time=2000 ms. T2-weight fluid-attenuated inversion recovery (FLAIR) images were acquired with a three-dimensional acquisition at a spatial resolution of 1.05×1×1 mm^3^. T1 images were post-processed in conjunction with T2-FLAIR images using FreeSurfer, generating cortical thickness and grey matter volume IDPs. Given research demonstrating improved accuracy of subcortical segmentation using FIRST compared with FreeSurfer^97^, subcortical grey matter volume IDPs from images post-processed with FIRST were also included. White matter macrostructural measures were also quantified from T1 and T2 FLAIR data using the BIANCA tool, generating WMH volume IDPs. Diffusion-weight images were acquired with a spatial resolution of 2×2×2 mm^3^ using two non-zero diffusion weightings (b=1000 and 2000 w/mm^2^). White matter microstructural measures were obtained from diffusion MRI data, including fractional anisotropy (DWI-FA), orientation dispersion index (DWI-ODI), intracellular volume fraction (DWI-ICVF), and free water (DWI-FW) IDPs. We selected 256 variables which had the most overlap with our validation cohorts. Individual datapoints were invalidated if (a) the value was 0 (not applied to WMH as 0 was a valid value for regional measures) or (b) an intracranial volume measurement was unavailable at the same instance (applied only to volumetric measures). Following data invalidation, if participants did not have (a) a complete cases neuroimaging dataset available for all 256 included variables (**Supplemental Table 2**) and (b) an intracranial volume measurement at that same instance, they were excluded from analyses. Prior to analysis, volumetric variables were corrected for intracranial volume. Grey matter volumetric measures were corrected using a full-sample residual approach as previously described by Wang et al.^98^ Given the skewed distribution of WMH data, WMH volumetric data were corrected using a ratio approach, as previously described by Wardlaw et al.,^99^ and then log-transformed. All neuroimaging data were scaled to z-scores prior to analysis.

VMAP neuroimaging data acquisition and post-processing have been previously described in detail^83^. For the current project, total grey matter volume was quantified from T1-weighted MRI data post-processed with Multi-Atlas. Brain MRI acquisition and processing in CARDIA has been reported previously.^91^ Grey matter volume was indexed to intracranial volume, white matter hyperintensity ratio was defined as the ratio between abnormal white matter volume and total white matter volume, hippocampus volume was defined as left plus right hippocampus volume indexed to intracranial volume.

### Cognitive assessment

In VMAP, participants completed a comprehensive neuropsychological protocol at enrollment and all follow-up visits (e.g., 18 months, 3 years, 5 years, 7 years, 9 years, 11 years). Cognitive composites for executive function and episodic memory were calculated using bifactor latent variable modeling, as previously described.^100^ The executive function composite leveraged item-level data from the Delis-Kaplan Executive Function System (D-KEFS) Color-Word Inhibition Test, D-KEFs Tower Test, Letter Fluency (FAS) Test, and D-KEFS Letter-Number Switching Test.^101^ The episodic memory composite leveraged item-level data from the California Verbal Learning Test, Second Edition and the Biber Figure Learning Test.^102^ The current project included MoCA score^103^ as a measure of global cognition along with both the episodic memory and executive function composite scores as cognitive outcomes of interest.

In CARDIA, cognitive assessments were performed at the Year 30 exam by trained study technicians. These include the Rey Auditory and Verbal Learning Test (RAVLT)^104^, the Digit Symbol Substitution Test (DSST)^105^, and Stroop Color and Word Test.^106^ RAVLT is a measure of verbal learning and memory, DSST is a measure of processing speed, executive function and working memory, and the Stroop Color and Word Test is a measure of executive function.

### Statistical methods

All UKB analyses were performed using R 4.4.0 on the UKB Research Analysis Platform (https://ukbiobank.dnanexus.com/).

#### Principal component analysis of neuroimaging data

Principal component analysis (PCA) was performed across the standardized neuroimaging dataset including 8,711 UKB participants without missing neuroimaging data using the *psych* R package.^107^ A scree plot of the first ten unrotated PCs was visually inspected, and the first three PCs were selected for varimax rotation. Outliers >5 standard deviations from the mean on any of the first three PCs were identified and removed (n=5), and the neuroimaging data was then re-standardized within the remaining sample (n=8,706). Following outlier removal, PCA was performed specifying 3 components and varimax rotation, and the final rotated components (PC1-PC3) and their corresponding loadings were extracted for downstream analyses.

#### Regression of neurostructural principal components against the blood proteome

Linear regressions separately related plasma protein measures to neurostructural PCs (one protein-PC pair per model). Base covariate models adjusted for age and sex. Fully adjusted models also included race, BMI, education, smoking status, TDI, and *APOE*-e4 status as covariates. Race was self-reported by participants at study entry and categorized for analysis as White, Black, Asian, Mixed, or Unknown/Other. Education was re-coded as a binary variable mirroring an approach shared by Davies et al.^108^ Smoking status was also re-coded as a binary variable, classifying participants as current or prior smokers versus never smokers. *APOE*-e4 carrier status was determined using single-nucleotide variant rs429358 and defined as either positive (e2/e4, e3/e4, e4/e4) or negative (e2/e2, e2/e3, e3/e3). False discovery rate correction was applied across models for each neurostructural PC using the Benjamini-Hochberg procedure to account for the number of plasma proteomic predictors. The threshold for significance was set *a priori* as an FDR-adjusted p-value <0.05.

#### Pathway enrichment analysis

Pathway enrichment analysis was performed using the R package *clusterProfiler*^109^ including four databases: Gene Ontology Biological Process (GO BP), Kyoto Encyclopedia of Genes and Genomes (KEGG), Reactome, and WikiPathways. Proteins were mapped from UniProt IDs to Entrez Gene IDs via R package org.Hs.eg*.db*.^110^ The background gene set was defined as all 2,907 unique proteins tested in UK Biobank from the Olink Explore 3072 platform. Redundant GO BP terms were removed using semantic similarity clustering included in the package. FDR<0.05 was used as the significance threshold throughout.

#### Anatomical localization of the neurostructural proteome with NeuroimaGene

The NeuroimaGene resource^111^ was leveraged to examine associations between GReX of genes corresponding to proteins identified in the UKBB base covariate linear regression models described above and neurostructural phenotypes. Proteins from base covariate linear regression models with FDR<0.05 were mapped to gene transcripts using HUGO gene nomenclature. The NeuroimaGene resource was used to extract associations between GReX and the IDPs associated with each PC, including FreeSurfer grey matter volumes and cortical thickness IDPs for PC1, DTI-FA and DTI-ICVF for PC2, and DTI-ISOVF and DTI-OD for PC3. All associations were corrected using a strict Bonferroni multiple testing correction with a threshold of 0.05. The mean normalized effect size magnitude of all significant GReX measures on each IDP was calculated. The aggregate effect size magnitudes for PC1 (grey matter volumes and cortical thickness) were visualized using the native plotting architecture in the NeuroimaGene R package. The Johns Hopkins University white matter atlas alongside the ggseg3d package were used to visualize the DTI results for PC2 and PC3.

#### Plasma multi-protein score derivation

To derive multi-protein scores for each neurostructural PC, all proteins significantly associated with each PC (FDR<0.05) in age and sex adjusted models were included as penalized factors for selection in LASSO models for each respective neurostructural PC (e.g., the 191 proteins related to PC1 at an FDR<0.05 were input as predictors in LASSO for PC1). Complete proteomic data, without imputation, was required for LASSO analyses, and all plasma proteomic datapoints were standardized to z-scores prior to analysis. LASSO models were implemented using glmnet via the *caret* package^112^ with α fixed at 1 and the optimal regularization parameter (λ) selected via 10-fold cross validation. For each PC, a multi-protein score was calculated as the weighted sum of all proteins with a non-zero LASSO coefficient. Multi-protein scores were also standardized to z-scores prior to further analysis. Model fit was assessed using Pearson’s correlation in both (a) the derivation sample used for LASSO analyses for each PC and (b) a separate internal validation sample of participants with neuroimaging data who were not included in LASSO due to missing proteomic data on the proteins input to LASSO models but had complete data on the LASSO selected proteins (e.g., a participant with missing data on the 191 proteins related to PC1 at an FDR<0.05 but with complete data on the 84 proteins selected by LASSO). Sample sizes for each LASSO model in UKB is as follows: PC1 derivation sample N = 1,647, validation sample N = 717; PC2 derivation sample N = 2,104, validation sample = 260; PC3 derivation sample N = 1,370, validation sample N = 994.

To use the multi-protein scores in GNPC and ADRC required translating the score from the Olink Explore 3072 platform (UKB) to SomaScan 7k (GNPC, ADRC). Using UniProt identifiers to match between the 2 platforms, we identified the relevant Somalogic aptamers for score construction. In the cases where >1 aptamer linked to 1 UniProt identifier, we selected the aptamer with the highest correlation between Olink and SomaScan using published data.^93^ In cases where no aptamer matched on UniProt identifier, that coefficient was removed resulting in truncated scores (PC1: 69 of 84; PC2: 68 of 102; PC3: 102 of 137), which demonstrated excellent calibration with the original scores in UK Biobank using Pearson’s correlation.

#### Survival analysis with multi-protein scores predicting incident dementia

UKB participants with complete data on the LASSO selected proteins (no requirement to have brain MRI data) were included in survival models for dementia. Dementia outcomes and time-to-event data were defined by UKB (Data Fields 42018, 42020, 42022, 42024). Participants with prevalent cases of dementia were excluded. Given only 1 participant died prior to the event/censor date, we opted to exclude that participant giving a sample size of 9,973 UKB participants included in survival analyses. Survival models were adjusted for age, sex, race, BMI, education, smoking status, TDI, *APOE*-e4 status, systolic blood pressure, high-density lipoprotein, and hemoglobin A1c. We compared performance of survival models using the multi-protein scores to models without the protein scores, evaluating C-statistics and net reclassification index (NRI; calculated at the 75^th^ percentile for NRI for events). To test for effect modification by age, sex, *APOE*-e4 status, and standard polygenic risk score (PRS) of Alzheimer’s disease (Data Field 26206), Cox models included interaction terms between the multi-protein scores and the modifying variable. Separate models were created for each combination of multi-protein score, effect modifier (age, sex, *APOE*-e4 status, PRS of AD), and set of adjustment covariates. Observing no significant interaction between the multi-protein scores and PRS of AD, we built Cox models including both terms, without the interaction term, and estimated the hazard ratios across quartiles of PRS and protein scores with the *emmeans* package.

In the Knight ADRC Cohort, AD was diagnosed at annual visits with study investigators. As previously mentioned, participants included had a final diagnosis of either cognitively normal or AD; other cognitive disorders (PD, ALS, etc.) were excluded. Time to event was defined as time to AD diagnosis or last follow-up. Models included adjustments for age, sex, race, education, smoking status, BMI, *APOE*-e4 status, and pTau217.

#### Proteomic differences across cognitive phenotypes (GNPC & VMAP)

To evaluate whether baseline plasma protein levels or multi-protein scores differed between GNPC disease groups (i.e., AD, ALS, FTD, or PD vs. cognitively normal reference group), linear regressions assessed the associations of disease (vs. cognitively normal) with plasma protein levels. Models were adjusted for age, sex, and contributing site. FDR correction was performed using the Benjamini-Hochberg procedure.

To evaluate whether baseline plasma protein levels or multi-protein scores differed between CU and MCI VMAP participants, analysis of covariance was performed. VMAP participants with early MCI were excluded from differential expression analyses due to small sample size (n=10). Base covariate models adjusted for age, sex, and cohort (i.e., Legacy, Expansion). Fully adjusted models also included race/ethnicity, BMI, education, smoking status, area deprivation index (ADI)^113^, and *APOE*-e4 status as additional covariates. FDR correction was performed using the Benjamini-Hochberg procedure.

#### Associations between multi-protein scores and longitudinal clinical trajectory (VMAP & CARDIA)

Linear mixed-effects regression models related baseline multi-protein scores to longitudinal clinical outcomes in VMAP (one score-outcome pair per model), including the interaction between follow-up time and multi-protein score as the term of interest. Outcomes of interest included CDR Sum of Boxes score, Montreal Cognitive Assessment (MoCA) total score, and total grey matter volume. Models with total grey matter volume were adjusted for total intracranial volume. Random effects for follow-up time and random intercepts were included in the model. Base covariate models adjusted for age, sex, and cohort, and fully adjusted models included race/ethnicity, BMI, education, smoking status, ADI, and *APOE*-e4 status as additional covariates. Model covariates were captured at baseline. FDR correction was performed using the Benjamini-Hochberg procedure.

In CARDIA, we leveraged extant proteomic data from Exam Years 15, 20, 25, and 30 with cognitive and brain MRI phenotypes measured at Year 30 and biomarkers of AD (pTau217, Aβ42-to-Aβ40 ratio [Aβ42/40], pTau217-to-Aβ42 ratio [pTau217/Aβ42]) measured at Year 35. To relate the Year 15 multi-protein risk score with the outcomes at Years 30 and 35, we used linear regression models adjusted for Year 30 age, sex, race, education and *APOE*-e4 status. To estimate the trajectory of multi-protein risk scores over time, we used a linear mixed effects regression model (restricting the model to participants with >1 timepoint), with a random intercept and slope per participant, for each protein score as a function of time. Participants’ slopes were extracted from the model, then used as predictors in linear regression models of cognitive, brain MRI, and AD biomarkers in combination with the Year 15 multi-protein risk score. FDR correction was performed using the Benjamini-Hochberg procedure. We further evaluated for effect modification between the Year 35 biomarker status and multi-protein risk score (Years 15-30) on Year 30 outcomes. In linear models, we sequentially included Year 35 biomarkers, Year 15 multi-protein score, estimated slope of multi-protein scores from Year 15 to 30, and included interaction terms between biomarkers and Year 15 multi-protein score, and interaction terms between biomarkers and slope. The following transformations were performed prior to standardization (z-score): Stroop, log(x); AD biomarkers (pTau-217, Aβ42/40, pTau-217/Aβ42), log(x+0.01); white matter hyperintensity ratio, log(x+ ½ min of non-zero values).

#### Study samples for single-nucleus transcriptome brain atlas and pre-processing

For scientific rigor and to facilitate reproducibility, selected methods on study samples and single-nucleus RNA-seq pre-processing are reproduced from previous work, with appropriate attribution provided herein. All subjects were volunteers enrolled in the Arizona Study of Aging and Neurodegenerative Disorders (AZSAND), a longitudinal study established in 1996 in Sun City, Arizona, investigating aging, cognition, and movement disorders in older adults. Brain and body autopsies were conducted through the Banner Sun Health Research Institute Brain and Body Donation Program (BBDP). Transcription levels in 101 superior frontal gyrus samples were generated by aligning sequence reads to the reference genome (GRCh38.p5) using Cell Ranger (10x Genomics) with default parameters and the −-include-introns option enabled. Preprocessing was performed using the standard Seurat (v4.0) workflow on each sample, as previously described.^114^ Briefly, nuclei with fewer than 200 detected genes or with >5% mitochondrial, >5% ribosomal, or >0.1% hemoglobin counts were excluded. Nuclei with abnormally high or low total UMI counts were also filtered out by fitting a sigmoid function to the distribution of log10-transformed UMI counts and defining the cutoff as ln(4) × the scale factor from the inflection point. Nuclei with a log10GenesPerUMI value <0.8 were additionally excluded. Doublets were identified using DoubletFinder (v3.0) and removed. Each Seurat object was normalized using SCTransform. During normalization, the number of genes, number of UMIs, percentage of mitochondrial genes, and cell-cycle scores were regressed out. Principal component (PC) analysis was performed using the first 3,000 variable genes. Reciprocal PCA (RPCA) was performed using the first 30 principal components (PCs) derived from the 3,000 most variable genes to identify anchors prior to integration with SCTransform. Cell clustering was carried out using the default Louvain algorithm with a resolution parameter of 0.5. Cell clusters were annotated using previously published snRNA-seq data (syn18485175) as a reference.

#### Sample and variant-level QC

Four participants were excluded, two because of missing sample identifiers and two because of missing metadata. Genotype quality control was then conducted on the remaining 99 participants using PLINK v1.90b5.2. SNPs and samples with call rates < 98% were removed, and only autosomal SNPs were retained. SNPs with MAF < 5% or Hardy-Weinberg equilibrium (HWE) P < 1×10^−6^ were excluded. Two heterozygosity outliers were removed. Relatedness was assessed using pairwise identity-by-descent estimates calculated in PLINK, and one participant from a related pair with PI_HAT > 0.2 was removed. Genetic ancestry for study participants was determined by using PLINK 2.0 alpha. The PCs of the study participants were compared to 1000 Genome phase 3 participants. After these quality control steps, 96 study participants were retained, 91 of whom were determined to be of European ancestry. After all quality control analyses, there was a total of 5,656,874 variants used in downstream analyses.

#### Development of models of genetically-regulated expression for single-nucleus transcriptome-wide association studies

Transcriptome-wide association studies (TWAS) integrate genetic variation with gene-expression information to identify genes whose genetically regulated expression (GReX) is associated with a complex trait or disease. Traditional TWAS typically relies on bulk-tissue expression reference panels, which average expression across many cell types and can therefore obscure cell type-specific regulatory effects.^115,116^ Here we leveraged single-nucleus RNA-seq data in brain to model GReX at the level of individual cell types. This single-nucleus resolution-enhanced TWAS (resTWAS) approach can help identify not only which genes are associated with a phenotype (AD), but also in which cells these genes may exert their effects on disease.

In conventional TWAS, for a given gene *g*, the expression in a tissue is

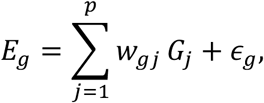

where *G*_j_ is genotype at SNP +, *w*_gj_ is the effect of the SNP on expression, *p* is the number of feature genetic variants, *E*_g_ is gene expression, and (_g_ is the non-genetic component. TWAS model building estimates the weights *w*_gj_ such as using penalized regression from a reference resource, as in our previous work.^115,116^ The genetically defined GReX is then estimated as follows

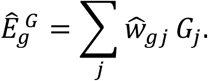

(Here, the superscript *G* denotes the cis-regulatory architecture, wherein the SNP features are obtained from the cis-region of the target gene.) The primary TWAS question is whether GReX shows a statistically significant relationship

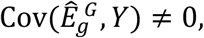

with the phenotype *Y*.

In resTWAS, expression is indexed not just by the gene *g*, but by the cell type *c*

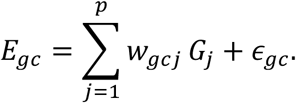

Thus, each gene can have a different regulatory architecture in different cell subpopulations

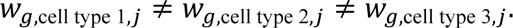

Thus, GReX is estimated in a cell type dependent manner

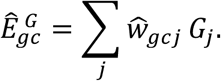

The weights are estimated as follows

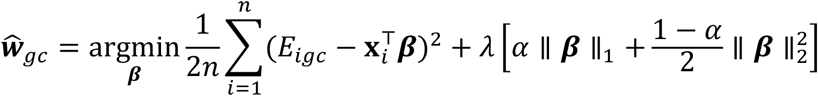

where *α* = 0.50 and sample *i*. Note the weighted sum of L1 and L2 penalties. In the phenotype model

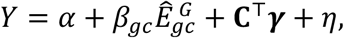

where **C** contains relevant (GWAS) covariates, the association being tested is therefore

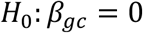

versus

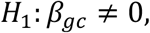

In single-cell or single-nucleus RNA-seq, we do not directly observe the continuous expression *E*_g*c*_. Rather, we observe *Y_i_*_g*c*_, i.e., the count for gene *g* in cell *i* of cell type *c*. Characteristically, the count *Y_i_*_g*c*_ = 0 is extremely common. Hence, we define the “measurement model.” We employ a model of count data using the zero-inflated negative binomial distribution.^117,118^ We introduce a latent variable

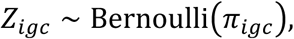

which defines whether the observation is from a zero or nonzero state, i.e., *Y_i_*_g*c*_ = 0 if *Z_i_*_g*c*_ = 1. If *Z_i_*_g*c*_ = 0, we model

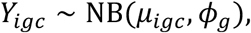

with *μ_i_*_g*c*_ and *φ*_g_ denoting the mean and dispersion parameters of the negative binomial (NB) distribution. We note

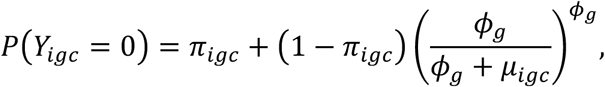

which shows that zeros are classified into structural (first term *π_i_*_g*c*_) versus sampling zeros. The mean is modeled as follows

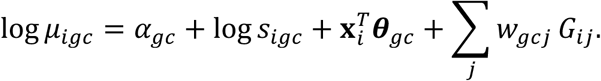

where *α*_g*c*_ is baseline expression, *s_i_*_g*c*_ is sequencing depth / library-size factor, and **x***_i_^T^* denotes the covariates. We leveraged pseudo-bulk gene expression, aggregating cells within cell types and donors and enabling inference at the donor level (versus individual cell level).

The GWAS (in this case, of AD and, separately, of Parkinson’s disease for trait-specificity comparison) provides a marginal association statistic *Z*_j_ for each SNP, while the single-nucleus reference panel of resTWAS models provides the expression weights *w*_g*c*j_. The TWAS statistic is

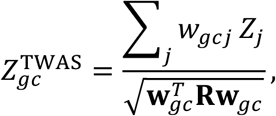

where

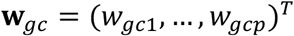

and **R** is the linkage disequilibrium (LD) correlation matrix among the SNP features. Under the null,

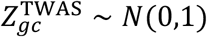

approximately, giving a gene-by-cell type p-value.

#### Validation of the neurostructural proteome associations with AD using single-nucleus brain atlas

We examined whether the neurostructural proteome was associated with the GReX of the corresponding genes in the brain using our single-nucleus brain atlas. We identified the proteins that could be mapped to their encoding genes and determined which cell types express the genes. For each such protein, we then tested the association of the corresponding GReX in a given cell type with AD using the resTWAS models (sample size: 111,326 clinically diagnosed/”proxy” AD cases and 677,663 controls), providing a genetically-anchored approach to evaluate whether genetic regulation of the gene in the cell type is associated with AD liability. Consider the sorted resTWAS p-values of the *K* proteins associated with the composite imaging phenotypes in a fixed cell type: *p_i_* where 1 ≤ *i* ≤ *K*. We compared the log_10_-transformed p-values with the expected quantiles 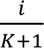 of the uniform null distribution

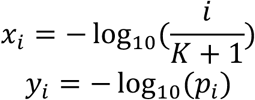

We tested for excess of small p-values from the cell type-dependent resTWAS associations with AD among these proteins

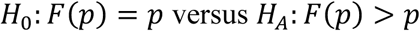

where *F* is the cumulative distribution function, using a one-sided KS test. Using a quantile-quantile (QQ) plot, we visualized the enrichment for association signals. We calculated the genomic inflation factor 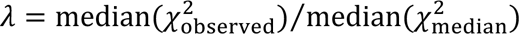. Finally, we compared the enrichment for AD resTWAS genetic signals among the AD neurostructural proteome with the level of enrichment for analogous genetic signals in Parkinson’s disease to test for trait specificity.^119^

## Supplemental Figures

**Supplemental Figure 1.**
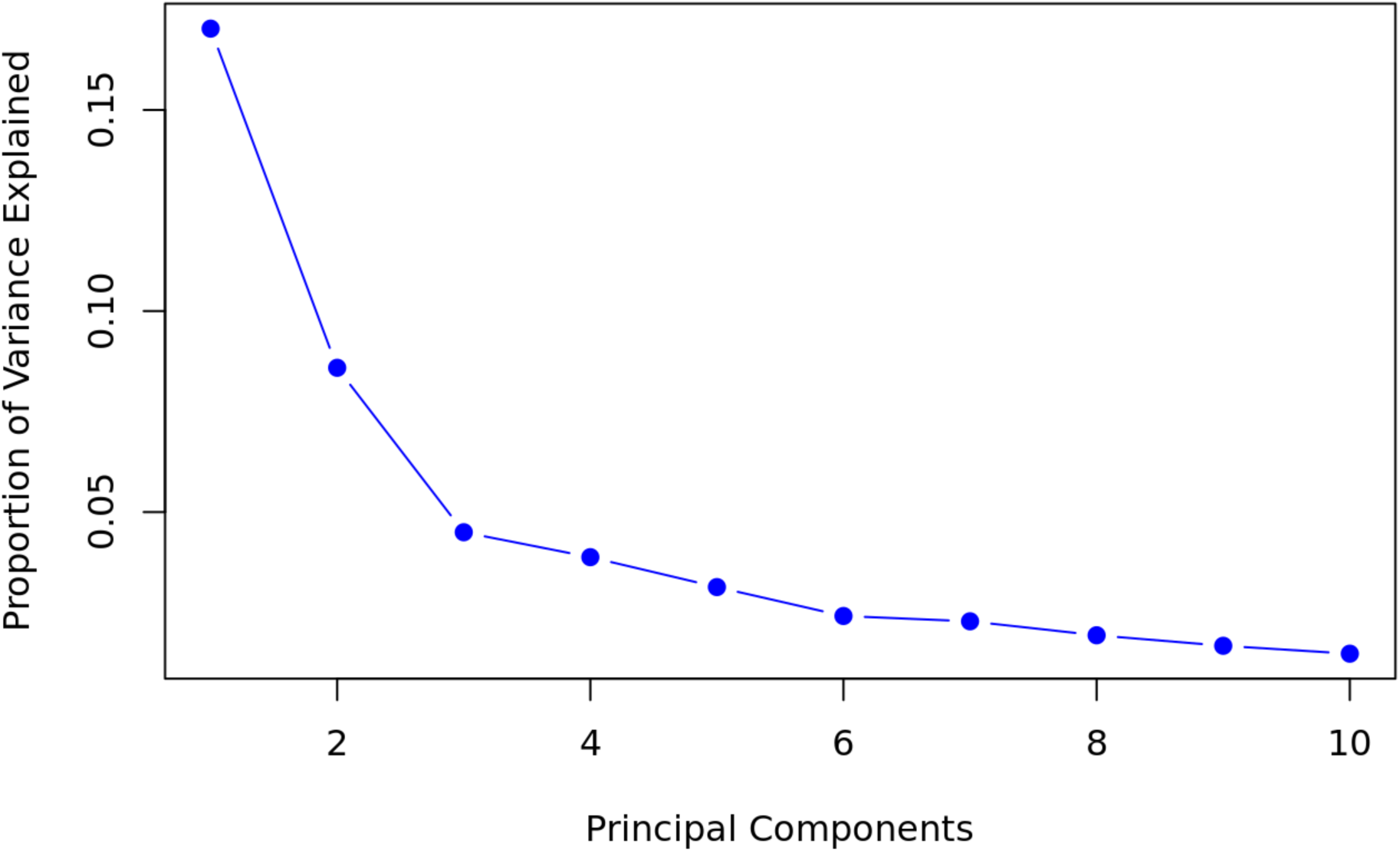
Principal component analysis of neuroimaging phenotypes. Scree plot of principal component analysis identifies 3 principal components.

**Supplemental Figure 2.**
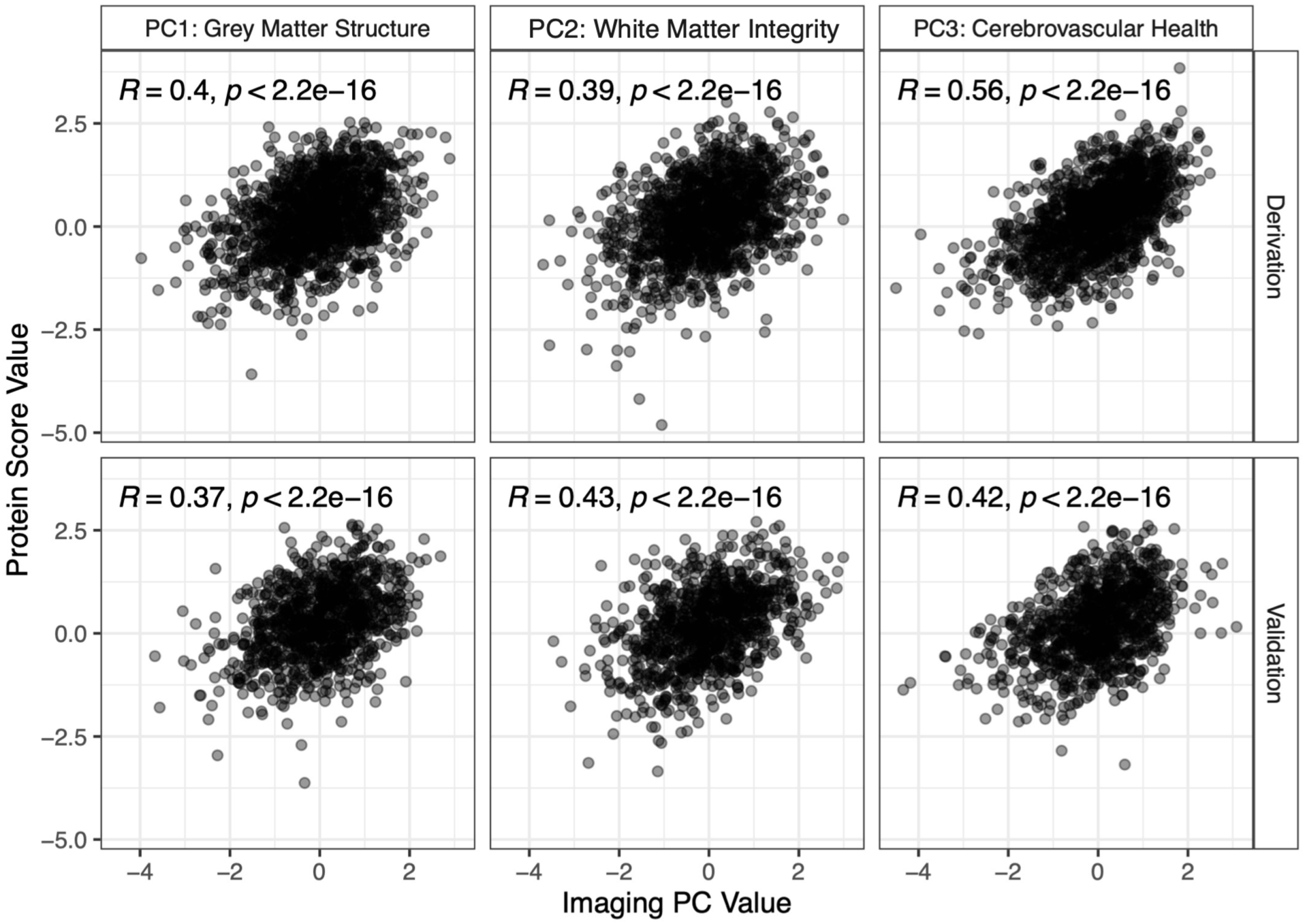
Model fit of multi-protein scores. Pearson correlation of the multi-protein scores (generated from LASSO model) with their corresponding neurostructural PCs. Sample sizes for each LASSO model in UKB is as follows: PC1 derivation sample N = 1,647, validation sample N = 717; PC2 derivation sample N = 2,104, validation sample = 260; PC3 derivation sample N = 1,370, validation sample N = 994.

**Supplemental Figure 3.**
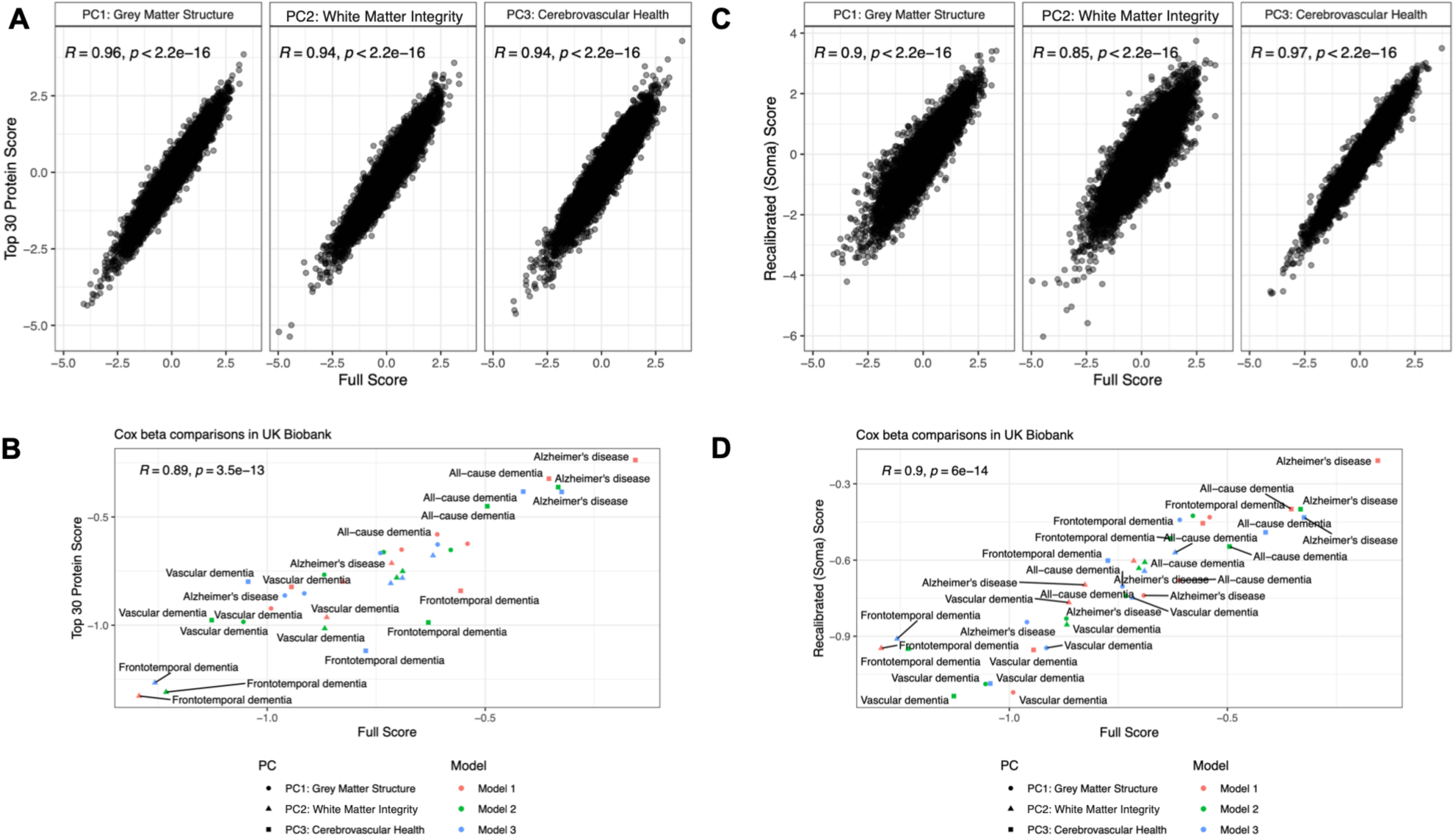
Performance of recalibrated multi-protein scores. **(A, C)** To test the performance of clinically translatable 30-protein scores and validate our findings in cohorts using Somalogic data, we recalibrated the scores as described in the Methods. Shown here is the correlation of recalibrated scores with the original scores using all LASSO selected proteins. **(B, D)** Comparison of Cox beta coefficients using the recalibrated scores to coefficients using the original scores using UK Biobank data. Model 1: age, sex; Model 2: Model 1 + race, body mass index, smoking status, education level, APOE-e4 status; Model 3: Model 2 + systolic blood pressure, high-density lipoprotein, hemoglobin A1c, Townsend Deprivation Index.

**Supplemental Figure 4.**
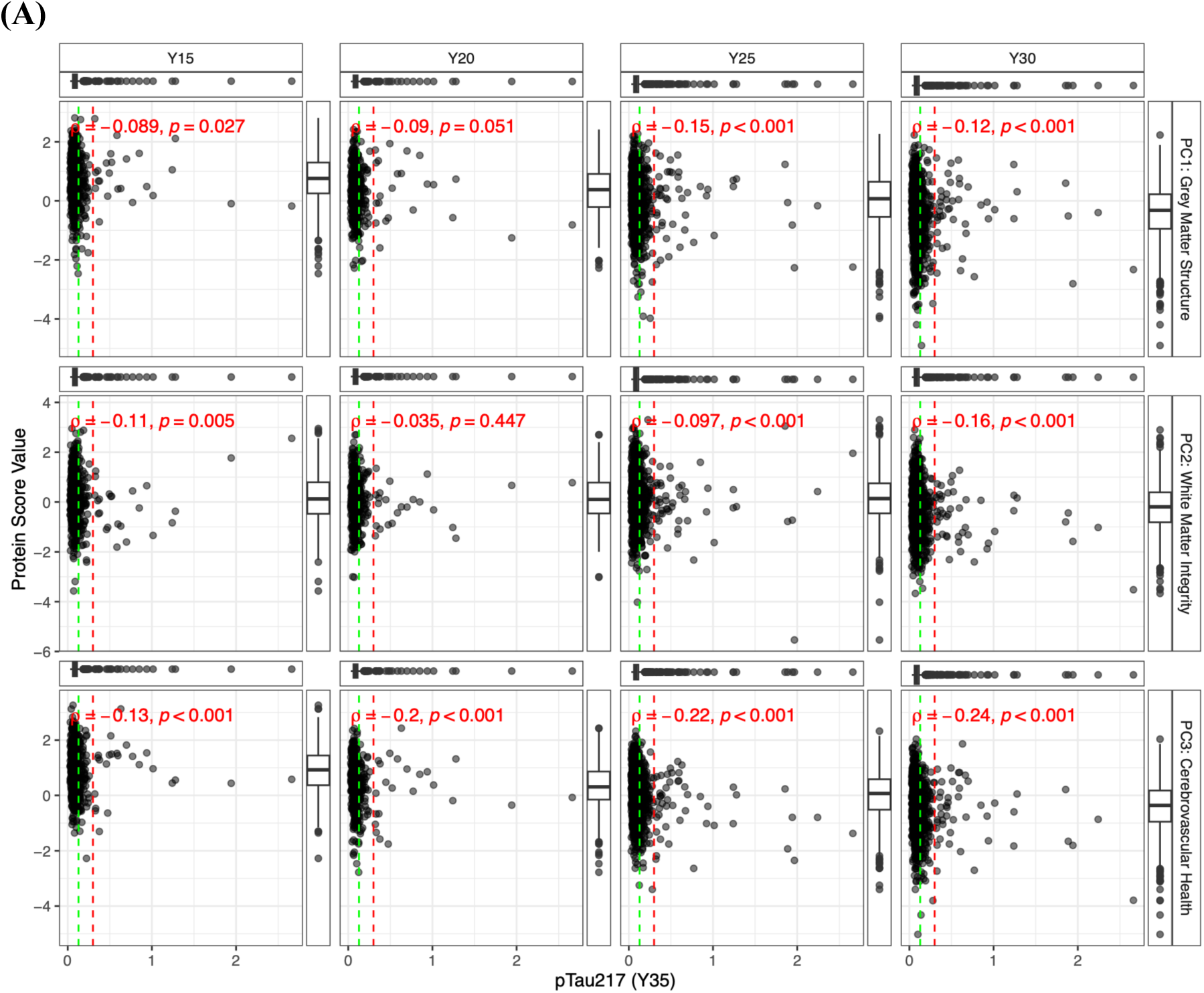
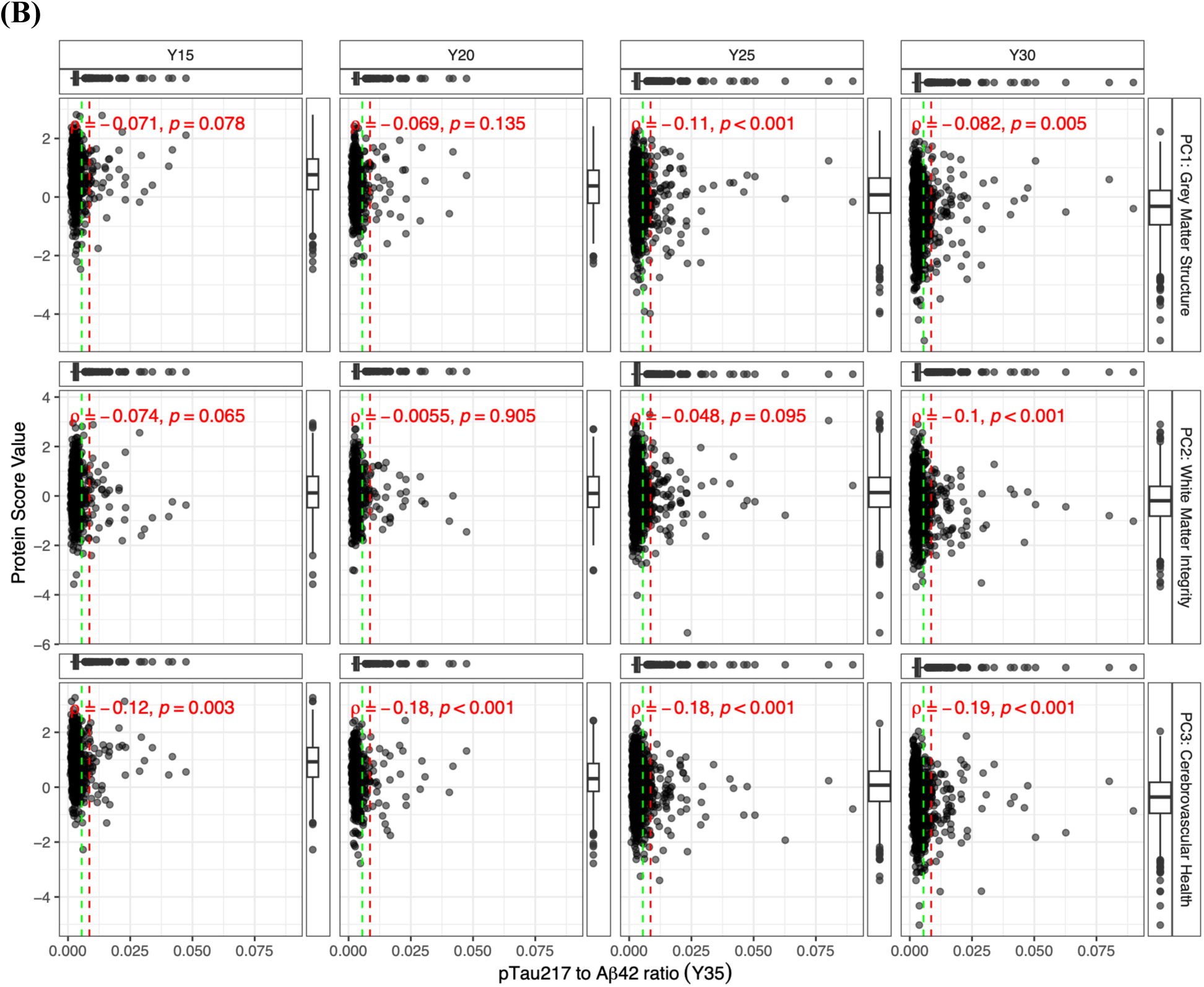
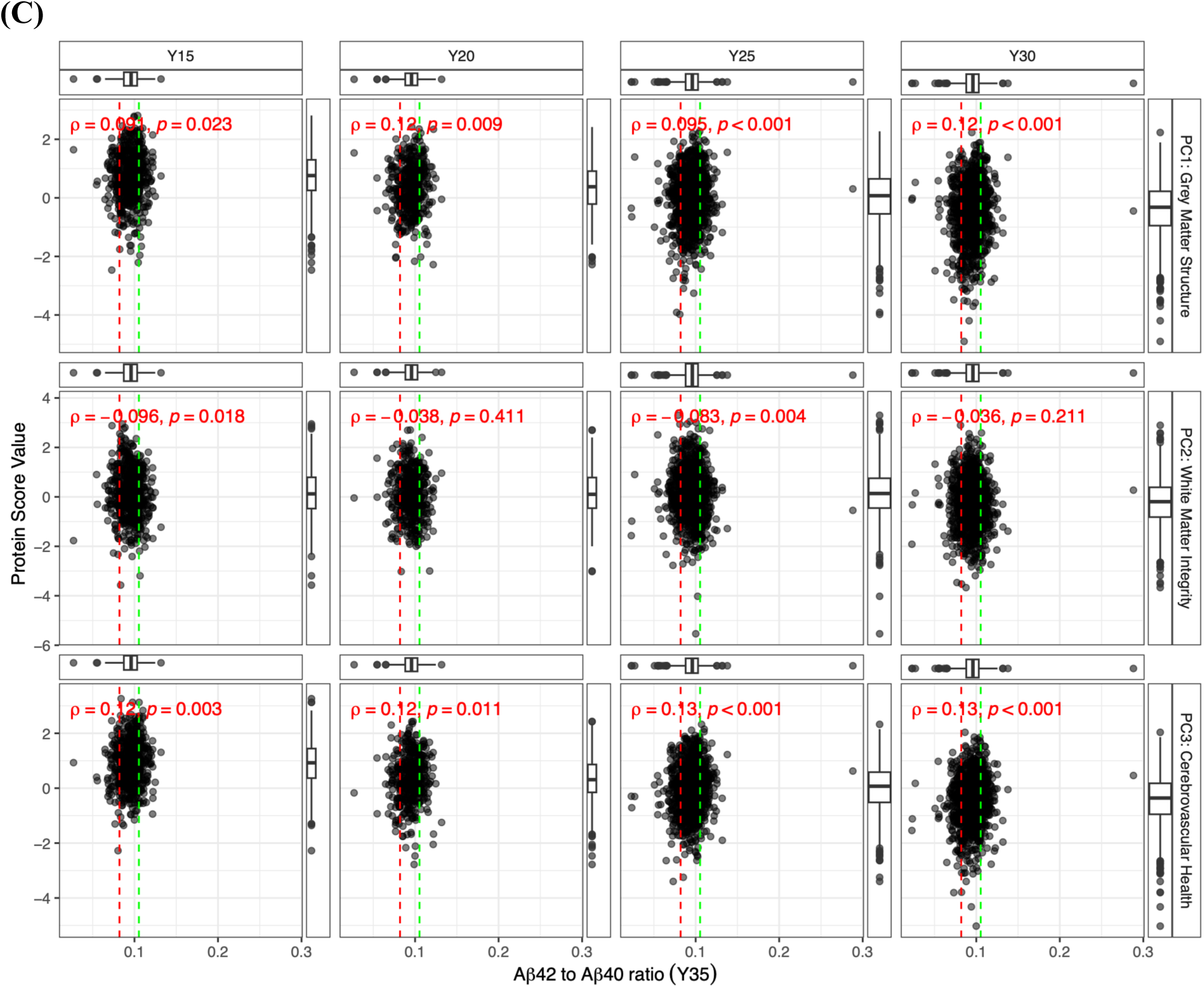
Correlation of AD biomarkers and multi-protein scores. **(A)** Spearman correlation plots of multi-protein scores at each CARDIA Exam (Y15-Y30) with established AD biomarkers. Box plots show distribution of variables along their respective axis. Red lines demarcate thresholds between intermediate and positive results. Green lines demarcate thresholds between negative and intermediate results. We used the following definitions for biomarker status: pTau217 − negative (≤0.128 pg/mL), intermediate (0.129–0.300 pg/mL), and positive (>0.300 pg/mL); pTau217/Aβ42 − negative (≤0.0055), intermediate (0.0056–0.0086), and positive (>0.0086); Aβ42/40 ratio − negative (≥0.1053), intermediate (0.0820–0.1052), and positive (<0.0820). Aβ=amyloid β. pTau217=phosphorylated tau217.

**Supplemental Figure 5.**
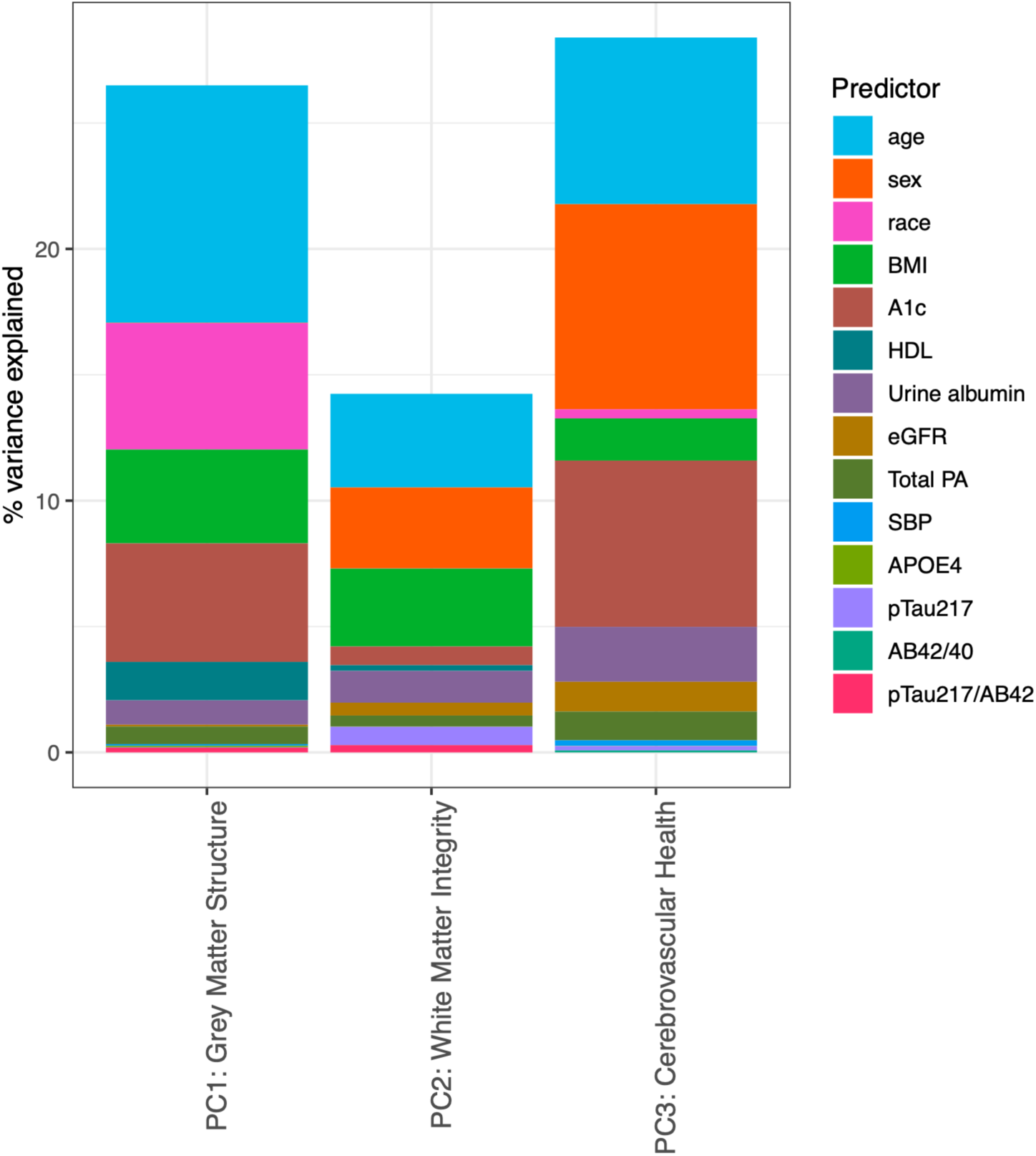
Explanatory components of multi-protein scores. Variance explained from Type 1 ANOVA models. Data for models came from CARDIA Exam Year 30, except the AD biomarkers (pTau217, Aβ, etc.) which were measured at Exam Year 35. BMI = body mass index; HDL = high density lipoprotein; eGFR = estimated glomerular filtration rate; PA = physical activity; SBP = systolic blood pressure; pTau217=phosphorylated tau217; AB = Aβ.

**Supplemental Figure 6.**
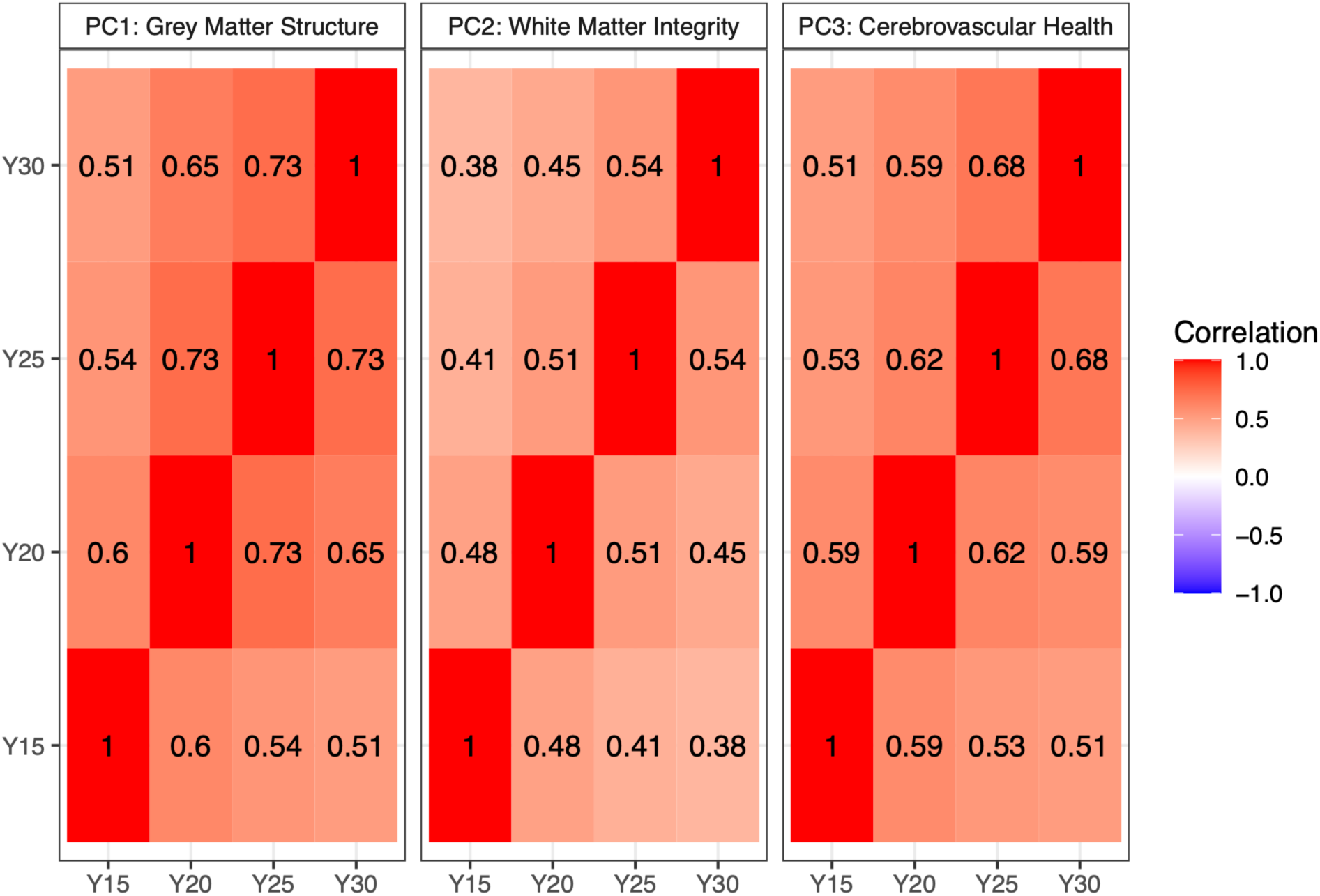
Correlation of multi-protein scores over time. Pearson correlation heatmap showing relatedness of multi-protein risk scores.

**Supplemental Figure 7.**
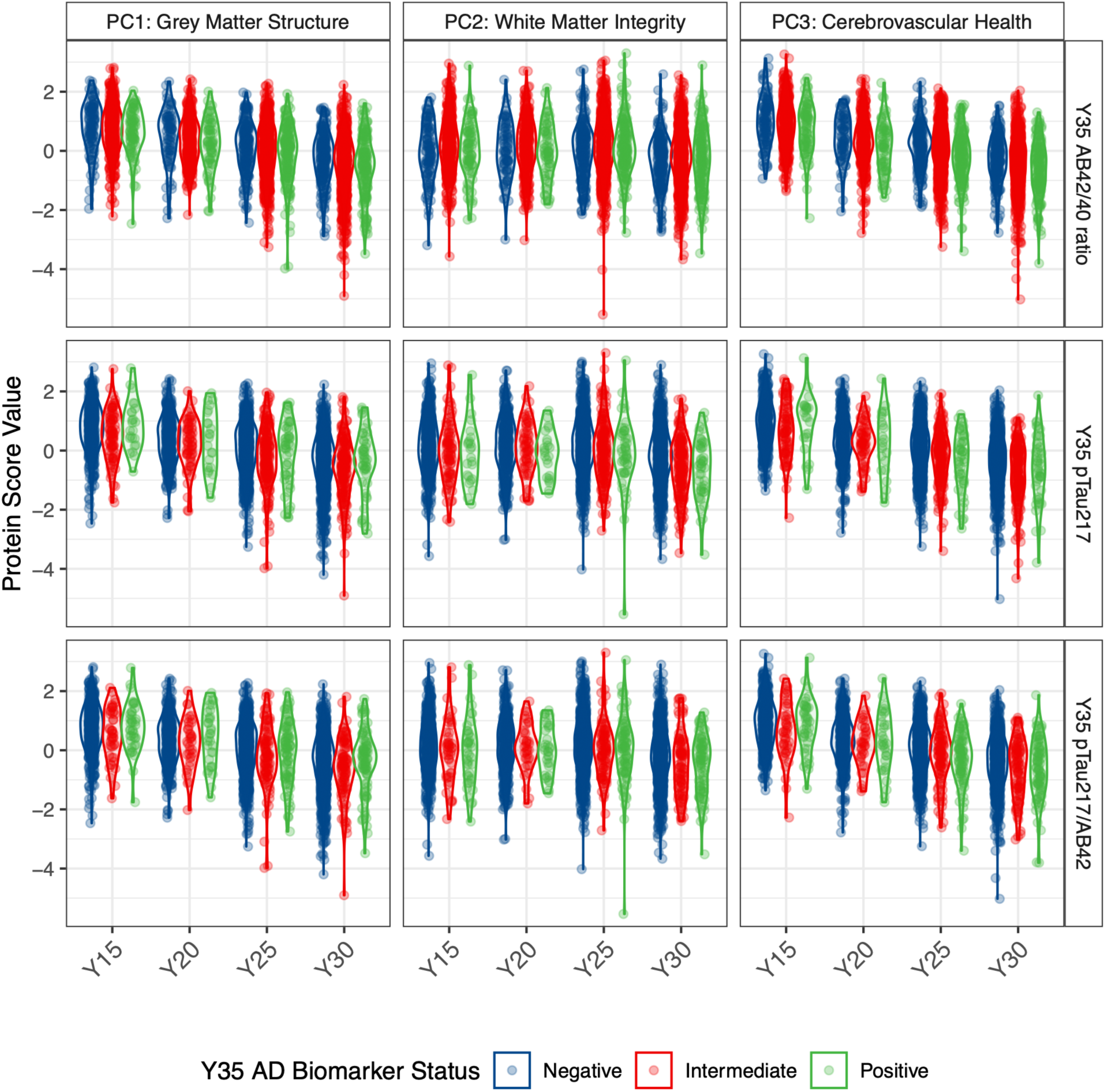
Longitudinal trends of multi-protein scores by AD biomarker status. Consistent with the overall trend of declining scores, we observed decline in multi-protein scores across all strata of AD biomarker statuses (biomarkers were measured at Year 35).

**Supplemental Figure 8.**
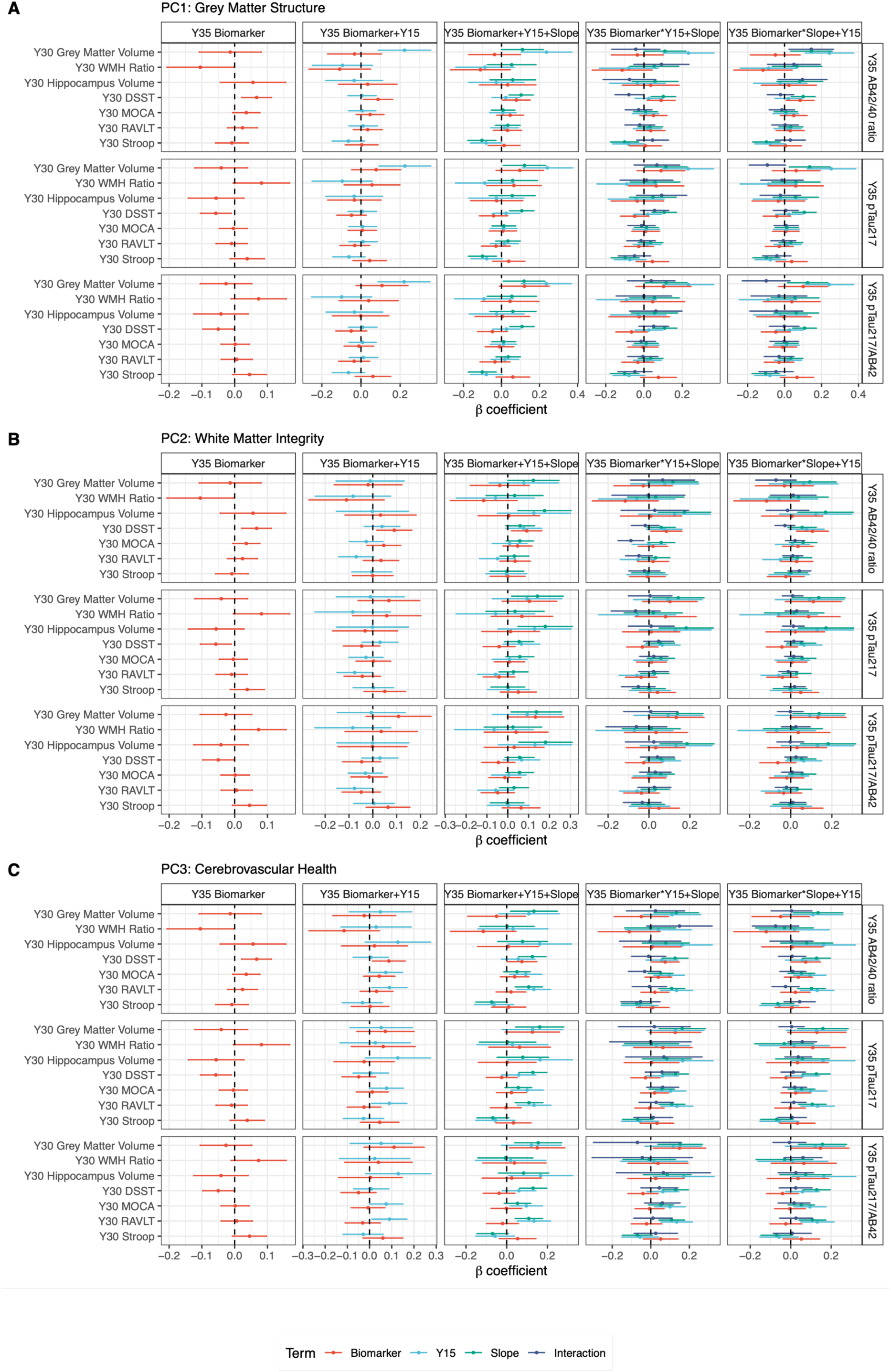
Interactive effects of AD biomarkers and multi-protein scores on cognitive function. Forest plots of linear models relating AD biomarkers (measured at Year 35) and multi-protein scores (measured over Years 15-30, with participant-level slopes estimated from mixed effects regression) with grey and white matter volume and cognitive function (assessed at Year 30). Overall biomarkers had weak relations with cognitive outcomes, which were reduced with inclusion of multi-protein scores. Y15 = multi-protein score at Year 15. Slope = participant-level slope estimated from mixed effects regression models. DSST = Digit Symbol Substitution Test. MOCA = Montreal Cognitive Assessment. RAVLT = Rey Auditory Verbal Learning Test; WMH Ratio = white matter hyperintensity ratio.

**Supplemental Figure 9.**
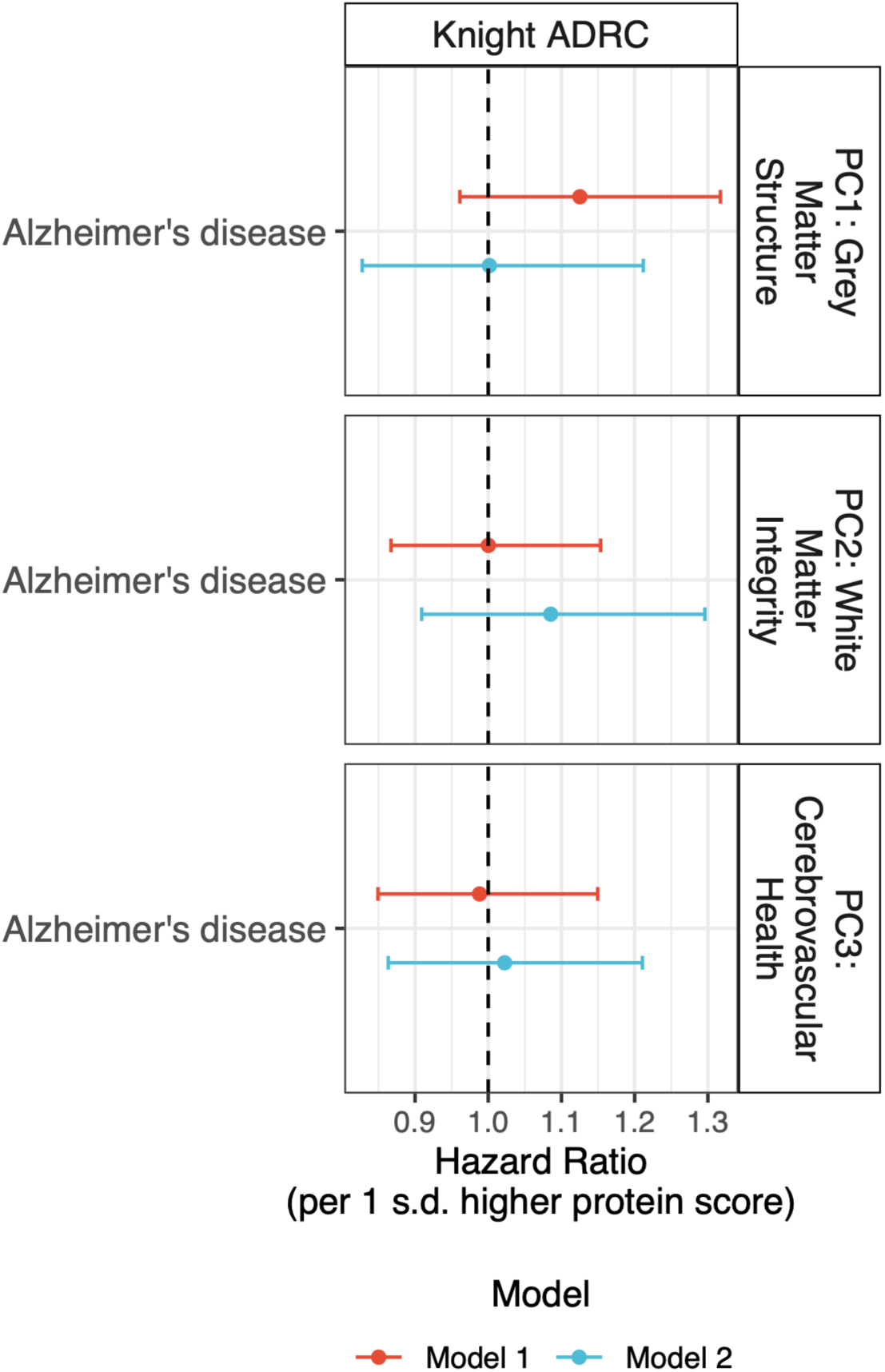
Performance of multi-protein scores in Knight ADRC. Forest plot of Cox models of Alzheimer’s disease in Knight ADRC. Model 1: age, sex. Model 2: Model 1 + race, education level, smoking status, BMI, and *APOE*-e4 carrier status. Error bars represent 95% confidence intervals of the hazard ratio.

**Supplemental Figure 10.**
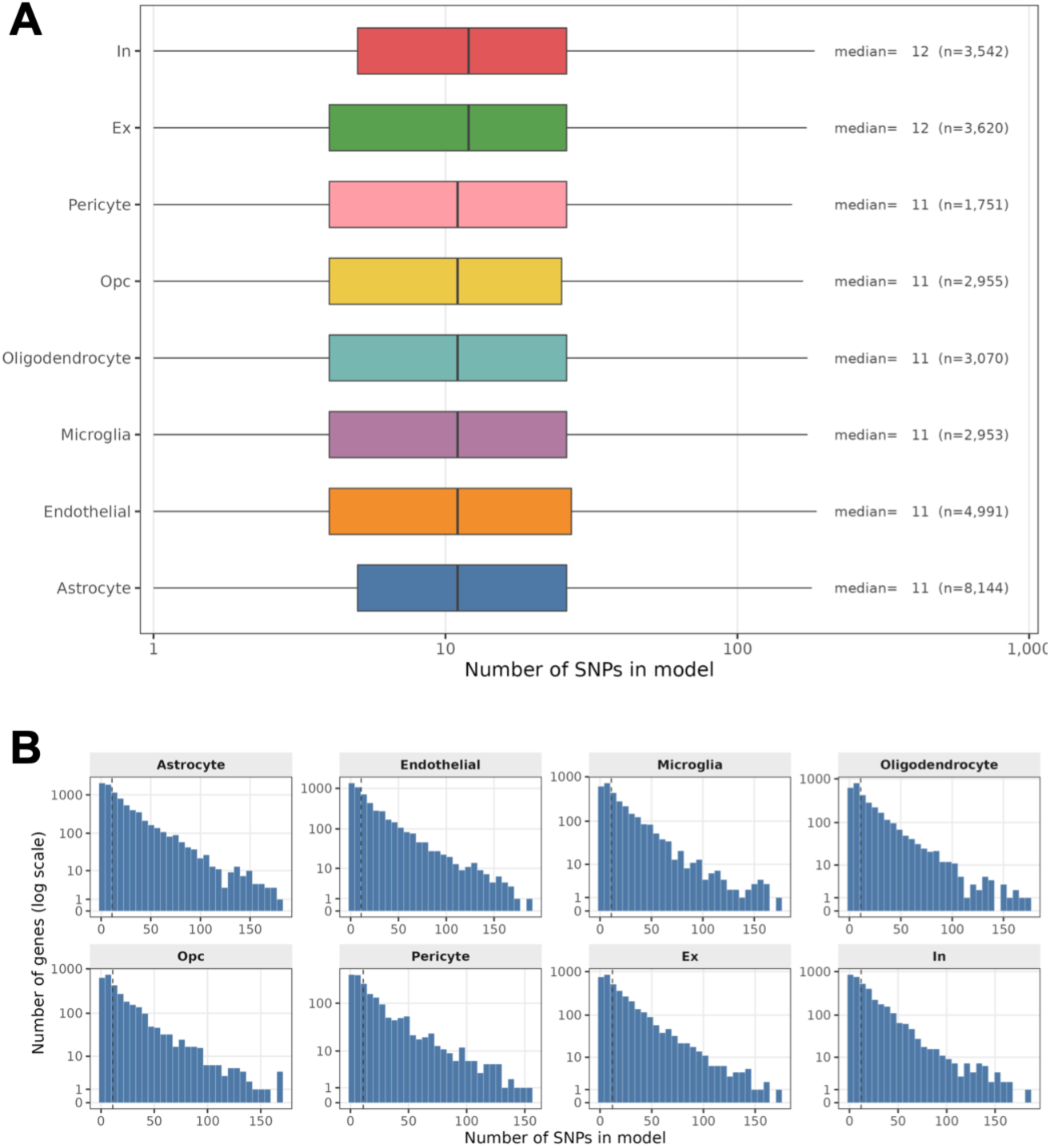
Complexity of gene expression models across cell types, indicating substantial allelic heterogeneity of *cis*-regulatory architecture. (A) We characterized the complexity of the resTWAS gene models. The median number of SNP features for optimal prediction was 11 across the major cell subpopulations except for the excitatory and inhibitory neurons, which had a median of 12. **(B)** We observed consistent allelic heterogeneity of gene expression in the cell types, with a large fraction of genes optimally predicted by >50 SNPs.

**Supplemental Figure 11.**
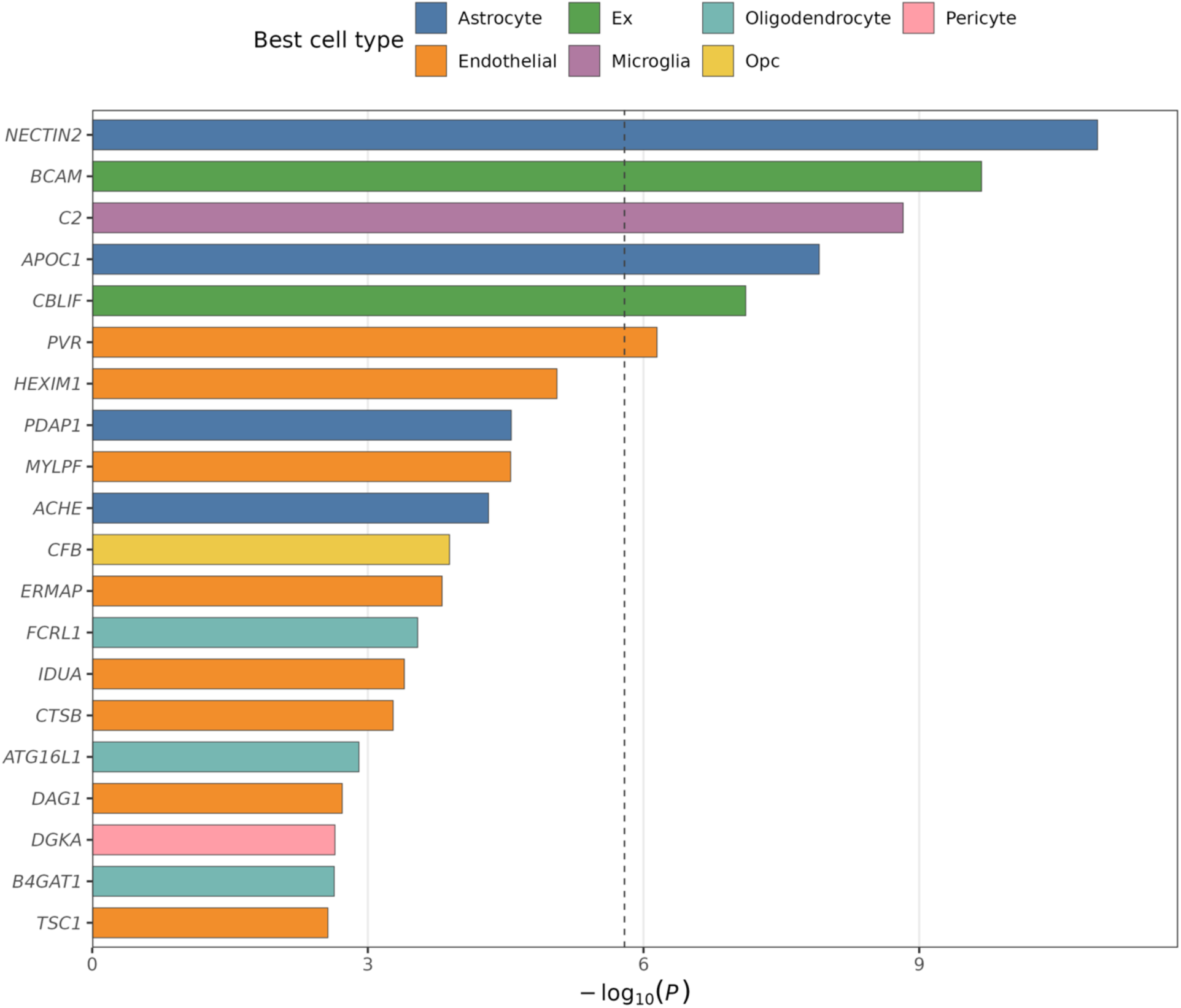
Top resTWAS genetic signals within the neurostructural proteome. Top findings from resTWAS analysis across the major cell types.

**Supplemental Figure 12.**
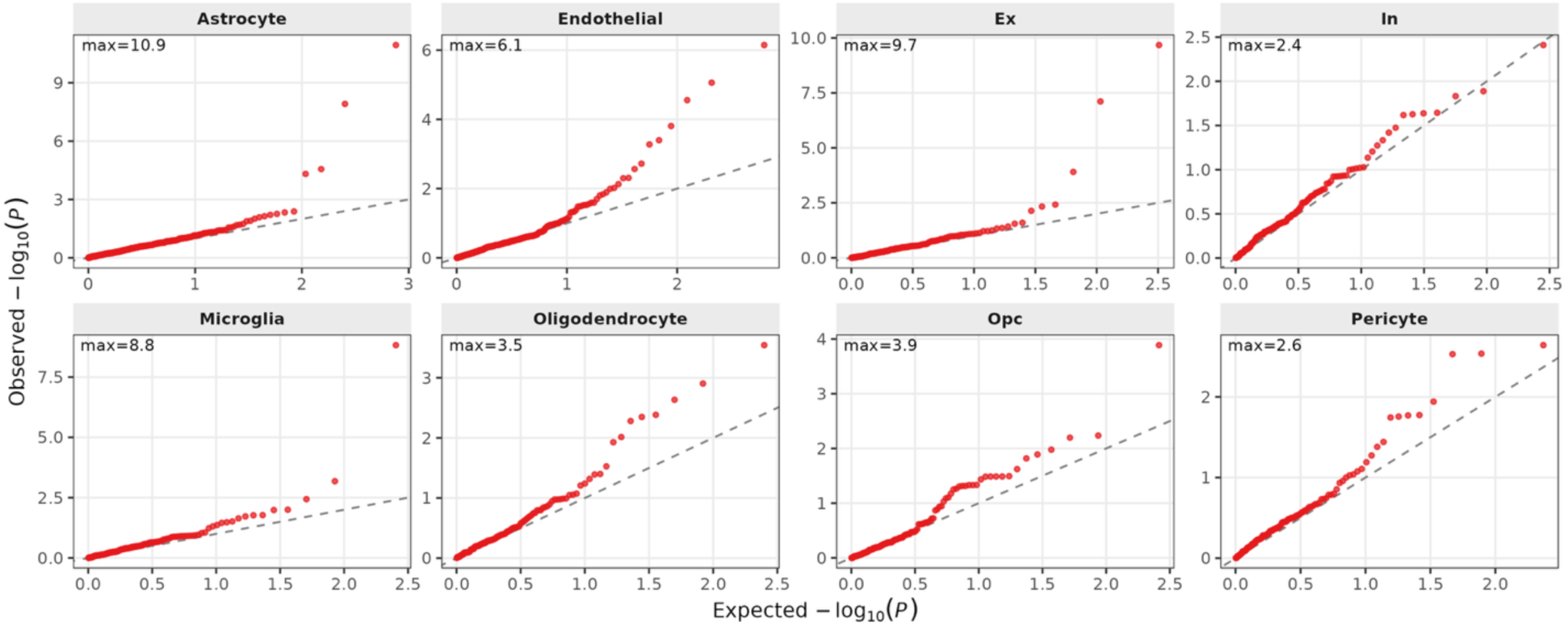
Cell-type dependent enrichment for resTWAS genetic signals among the genes encoding the neurostructural proteome. The neurostructural proteome showed significant cell type dependent enrichment for resTWAS associations with AD (as captured by leftward shift of the Q-Q plot) across the major cell subpopulations in the brain, except for inhibitory neurons.

## Notes

### Author Declarations

The Institutional Review Board at Vanderbilt University Medical Center approved this study.

